# An integrated multi-omics analysis of sleep-disordered breathing traits across multiple blood cell types

**DOI:** 10.1101/2022.07.09.22277444

**Authors:** Nuzulul Kurniansyah, Danielle A Wallace, Ying Zhang, Bing Yu, Brian Cade, Heming Wang, Heather M. Ochs-Balcom, Alexander P Reiner, Alberto R Ramos, Joshua D Smith, Jianwen Cai, Martha Daviglus, Phyllis C Zee, Robert Kaplan, Charles Kooperberg, Stephen S Rich, Jerome I Rotter, Sina A. Gharib, Susan Redline, Tamar Sofer

## Abstract

**Background:** Sleep Disordered Breathing (SDB) is characterized by repeated breathing reductions or cessations during sleep, often accompanied by oxyhemoglobin desaturation. How SDB affects the molecular environment is still poorly understood.

**Methods:** We studied the association of three SDB measures: the Apnea Hypopnea Index (AHI), average and minimum oxyhemoglobin saturation during sleep (AvgO2 and MinO2) with gene expression measured using RNA-seq in peripheral blood mononuclear cells (PBMCs), monocytes, and T-cells, in ∼500 individuals from the Multi-Ethnic Study of Atherosclerosis (MESA). We developed genetic instrumental variables (IVs) for the associated transcripts as polygenic risk scores (tPRS), then generalized and validated the tPRS in the Women’s Health Initiative (WHI). Next, we constructed the tPRS and studied their association with SDB measures (to identify potential reverse causal associations) and with serum metabolites (to identify downstream effects) in ∼12,000 and ∼4,000 participants, respectively, from the Hispanic Community Health Study/Study of Latinos (HCHS/SOL). Finally, we estimated the association of these SDB measures with transcript IV-associated metabolites in HCHS/SOL, to verify complete association pathways linking SDB, gene expression, and metabolites.

**Results:** Across the three leukocyte cell types, 96 gene transcripts were associated with at least one SDB exposure (False Discovery Rate (FDR) p-value <0.1). Across cell populations, estimated log-fold expression changes were similar between AHI and MinO2 (Spearman correlations>0.90), and less similar between AvgO2 and the other exposures. Eight and four associations had FDR p-value<0.05 when the analysis was not adjusted and adjusted to BMI, respectively. Associations include known genes that respond to (*PDGFC*) and regulate response to (*AJUBA*) hypoxia. We identified a complete “chain” linking AvgO2, *P2RX4*, and butyrylcarnitine (C4), suggesting that increased expression of the purinergic receptor *P2RX4* may improve average oxyhemoglobin saturation and decrease butyrylcarnitine (C4) levels.

**Conclusions:** Our results support a mechanistic role for purinergic signaling and hypoxic signaling, among others, in SDB. These findings show differential gene expression by blood cell type in relation to SDB traits and link *P2XR4* expression to influencing AvgO2 and butyrylcarnitine (C4) levels. Overall, we employed novel methods for integrating multi-omic data to evaluate biological mechanisms underlying multiple SDB traits.

## BACKGROUND

Sleep-disordered breathing (SDB) is a common disorder, affecting an estimated 24% of male and 9% of female adults in the U.S. (1). SDB is characterized by episodic periods of breathing cessations and reductions during sleep, often accompanied by oxyhemoglobin desaturation (2, 3), and is associated with cardiometabolic, vascular, and cognitive outcomes (4–7). SDB is also strongly associated with inflammation (8, 9). While obesity is a strong risk factor for SDB, SDB is also heritable independent of body mass index (BMI) (10, 11). The underlying molecular processes by which SDB affects health outcomes are still being studied (12), with interest in understanding SDB-related hypoxia during sleep on cardiometabolic and vascular measures in humans and in animal models (13–15).

In investigating the molecular changes caused by SDB, previous studies showed changes in distributions and activation of white blood cells (16–18) and inflammatory cytokines (19) in individuals with obstructive sleep apnea (OSA). Other studies reported changes in gene expression in white blood cells following treatment using continuous positive airway pressure (CPAP), or following CPAP withdrawal (20–23), supporting a causal role between SDB-related physiological stressors (such as hypoxia) and immune cell gene expression. Some studies, including those from our group, also reported cross-sectional transcriptomic association with SDB measures from observational studies (23, 24). However, these studies focused on a single cell population, and it is unknown whether and how transcriptional effects of SDB differ among circulating leukocyte subpopulations. Likewise, it is yet unknown how SDB-alterations in gene expression translate to metabolic changes. A few previous studies reported associations of blood metabolites with SDB phenotypes, independently of transcriptomics. For example, one study reported change in serum metabolite levels, evaluated on an untargeted platform that surveyed a few hundred metabolites, after multi-level sleep surgery (25). Most other studies considered specific, targeted metabolite changes in sleep disorders (see reviews in (26)).

Large, untargeted, omics surveys are now becoming available in cohort studies, providing an opportunity to study the association of SDB with well-defined, genetically-regulated molecular measures. We deploy a systems biology approach integrating genomic, transcriptomic, and metabolomic data to identify potential pathways in tissue-specific mechanisms driving SDB-related morbidity.

Utilizing data from the Multi-Ethnic Study of Atherosclerosis (MESA) and the Hispanic Community Health Study/Study of Latinos (HCHS/SOL), we examined multi-omics data to investigate signaling mechanisms underlying SDB traits. First, we used transcriptomics data measured in peripheral blood mononuclear cells (PBMCs), T-cells and monocytes, assayed by the Trans-Omics for Precision Medicine (TOPMed) program, to perform transcriptome-wide association study of SDB-related phenotypes (measured via overnight polysomnography) in MESA. We compared the results across different peripheral blood cell populations. With these data, we constructed transcript polygenic risk scores (tPRS) predicting transcript expression using genetic data. Next, we built these tPRS in the Women’s Health Initiative (WHI) and tested them for association and generalization with their transcripts in whole blood. We calculated the tPRS that generalized in HCHS/SOL. Finally, we applied these tPRS to SDB traits and metabolites in HCHS/SOL to investigate how SDB phenotypes potentially propagate via transcription to metabolic changes in serum, and on the other direction, to assess potential reverse association by which transcript expression causes changes in SDB phenotypes.

## METHODS

### Overall study design and purpose

The overall purpose of the study was to investigate the multi-omics signaling mechanisms underlying SDB traits to better understand possible drivers of morbidity in SDB. The study design and purpose of each analysis component is illustrated in **Figure 1**. Briefly, panel A demonstrates the set of associations investigated: SDB phenotypes lead to transcriptional changes which in turn lead to metabolic changes; panel B describes the analysis steps taken to study the potential chain of associations, and the goal of each of these steps. To optimize the available sample size and leverage the fact that transcription is, to some extent, genetically determined, we utilized two separate cohorts to identify the biological components associated sleep exposure to metabolomic changes. **Figure 2** further illustrates potential causal relationships underlying a set of measures, and the assumptions that we used to interrogate some of them. Thus, we first performed transcriptome-wide association studies for SDB phenotypes in MESA. For each transcript associated with a SDB trait (FDR p-value <0.1), we used genetic data to construct a transcript Polygenic Risk Score (tPRS) to serve as a predictor of the transcript. Next, to reduce false positive associations in subsequent analyses, we constructed these tPRS in the WHI and tested their association and generalization with their transcripts in whole blood. We proceeded with tPRS results that generalized (p-value <0.05), and constructed and tested them latter for association with SDB phenotypes in HCHS/SOL. If a tPRS was associated with the SDB phenotype in HCHS/SOL, it was interpreted as evidence of reverse association, i.e., the transcript may contribute to SDB. We then calculated the association of the tPRS with metabolites in HCHS/SOL. Lastly, we used another analytic step to support the existence of an association chain linking a sleep exposure, a transcript, and a metabolite: we required evidence of association between the sleep exposure and the metabolite in HCHS/SOL (i.e. any of the potential pathways in column A of **Figure 2**). If the tPRS was associated with the sleep exposure in HCHS/SOL, it lent support to association chains where the transcript affects both SDB and metabolite levels.

**Figure 1:**
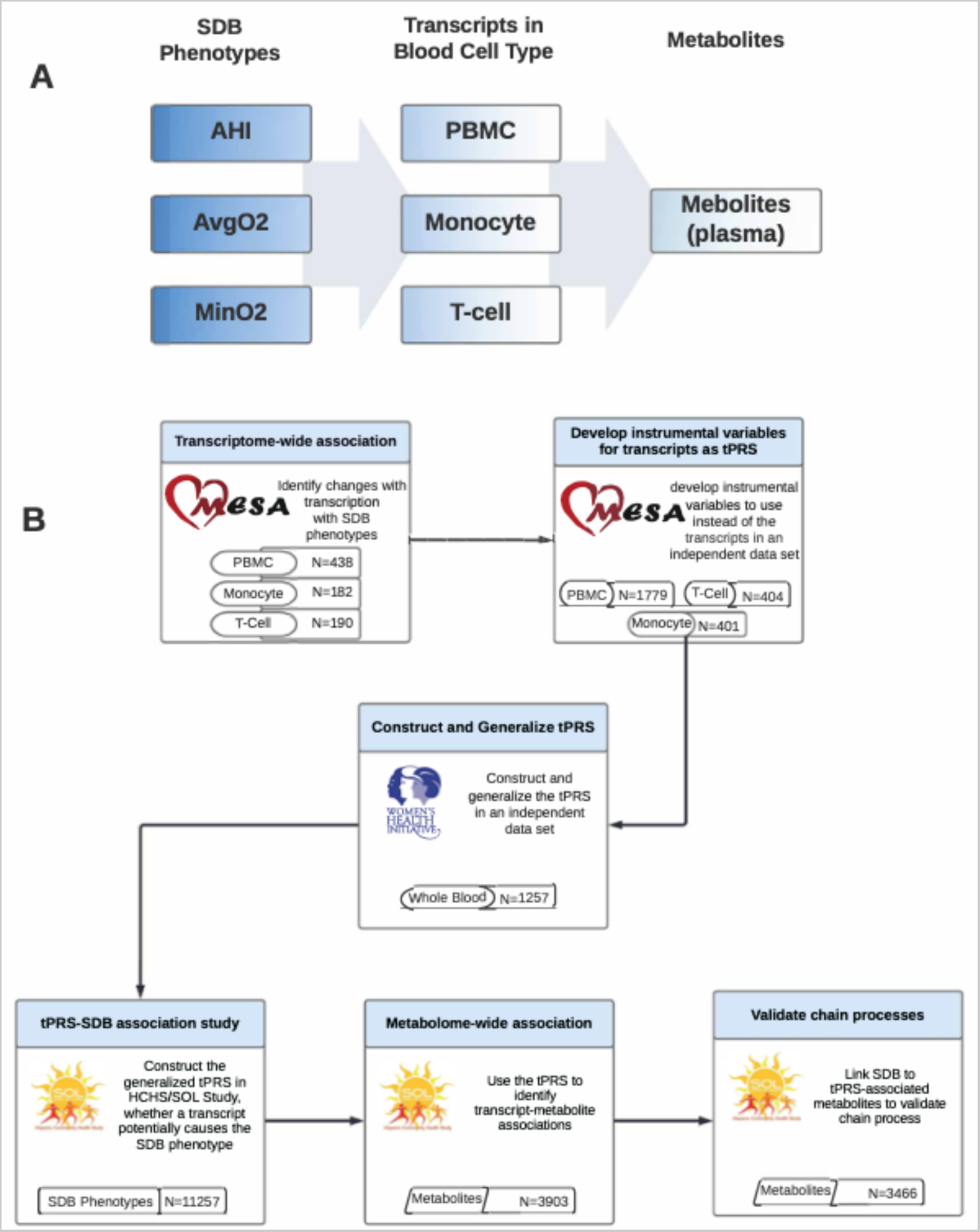
Overall study design of the reported analysis. Flow charts illustrating the methodology and purpose of the analysis. The chart portrays the three SDB phenotypes evaluated in the analysis, the three blood tissues with transcript expression measurement, and demonstrates each step of the analysis with associated goals, cohorts, and reasons why step was performed. Penal A: conceptual linking between SDB measures, transcript expression, and metabolites. Panel B: analytic steps supporting the study of the conceptual links. PBMC tPRS analysis sample sizes in MESA correspond to data from two visits (some individuals were used twice, appropriately accounted for by mixed models).

**Figure 2:**
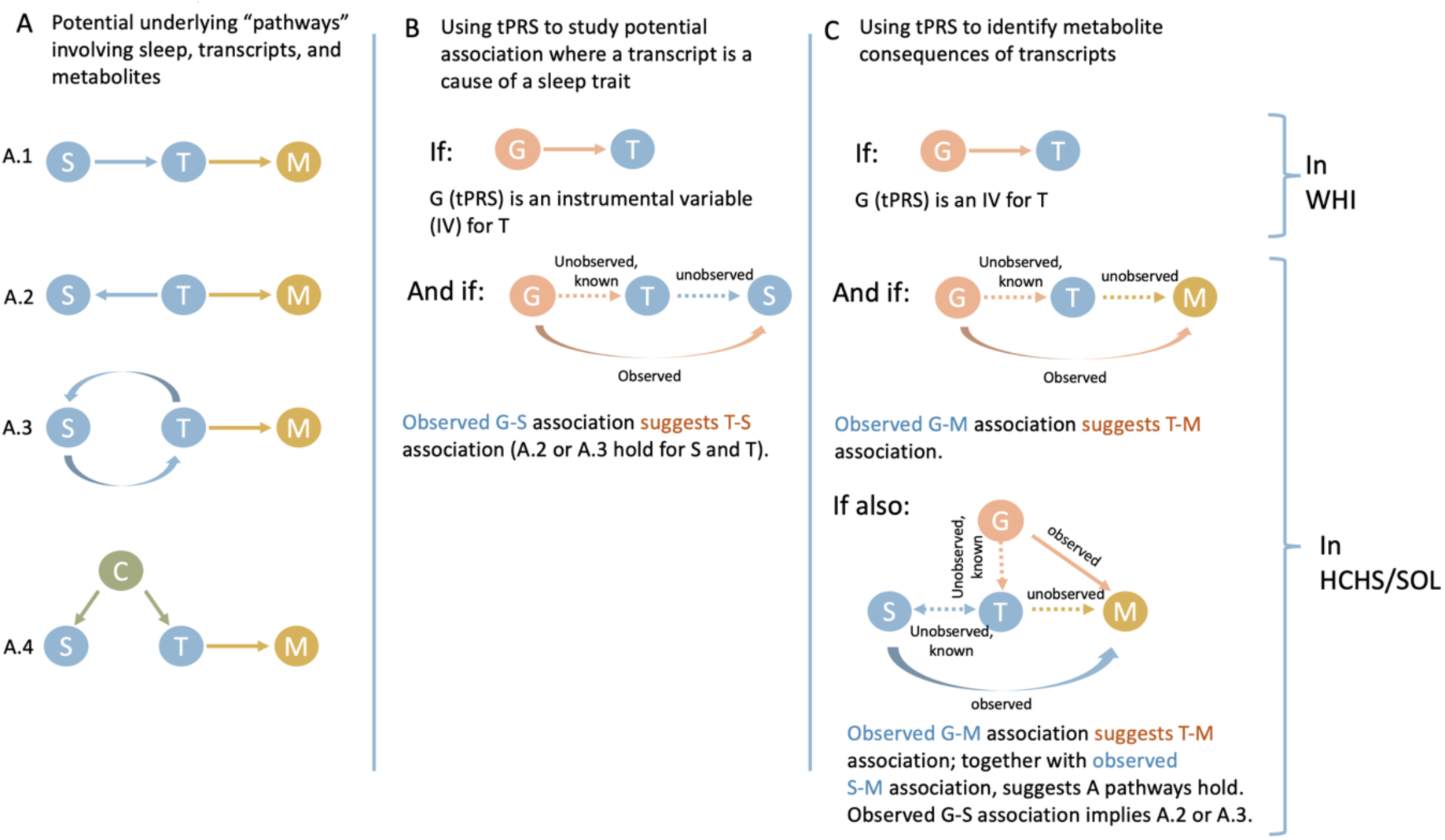
Potential association “chains” of SDB traits, transcript expression, and metabolites, addressed in this study. The figure illustrates association chains, or pathways, potentially linking an SDB trait, a transcript, and a metabolite. Here we assume that an association between a sleep trait and a transcript was detected in MESA and is assumed “known” for follow-up analysis in WHI in HCHS/SOL. Column A demonstrates potential forms of causal associations between the sleep trait and the transcript, including (A.4) the settings where an association exists due to a common cause, e.g. BMI. Our metabolomics analysis may only detect transcript-metabolite associations, i.e. any sleep-metabolite link is via transcript levels. Column B demonstrate a potential conclusion from an association between a tPRS, validated in WHI and used as an instrumental variable (IV) of a transcript, and a sleep trait: if an association is detected, it provides evidence that changes in transcript levels are upstream (a cause of) changes in sleep trait levels. Column C demonstrates potential conclusions from analyses linking tPRS and a sleep trait to a metabolite. A tPRS is used to link a transcript to a metabolite, and an association, if exists, is likely causal. A sleep-metabolite association should exist if the sleep-transcript and tPRS-transcript associations hold, and therefore observing such an association validates the existence of any of A pathways. Further association between the tPRS and the sleep trait narrows down the potential association chains to A.2 or A.3.

### Participating studies

As described in **Figure 1**, our analysis included three studies: MESA, WHI, and HCHS/SOL each contributing to different analytical steps. The three studies are described in the Supplementary Information. In brief, MESA, our primary study used for discovery of SDB-transcript associations, is a longitudinal cohort study (27). The 1^st^ and 5^th^ MESA exams took place between 2000-2002, and 2010-2012, respectively, and whole blood was drawn from participants in both exams. For about 1,000 participants, blood was used later for RNA extraction in at least one of the exams. In addition, a sleep study ancillary to MESA occurred shortly after MESA exam 5 during 2010-2013. Sleep study participants underwent single night in-home polysomnography, as previously described (28). The number of individuals with each type of data and at each time point (exam 1 and exam 5) varies. **Figure S1** in the Supplementary Information visualizes the data flow and overlaps across the various measures used in this study: whole-genome genotyping, RNA-seq, and sleep. All MESA participants provided written informed consent, and the study was approved by the Institutional Review Boards at The Lundquist Institute (formerly Los Angeles BioMedical Research Institute) at Harbor-UCLA Medical Center, University of Washington, Wake Forest School of Medicine, Northwestern University, University of Minnesota, Columbia University, and Johns Hopkins University.

The WHI was here used to identify tPRS that could be confidently used as IVs for their traits. It is a prospective national health study focused on identifying optimal strategies for preventing chronic diseases that are the major causes of death and disability in postmenopausal women (29). 11,071 WHI participants have whole-genome sequencing data via TOPMed, and 1,274 of these participants have RNA-seq measured in venous blood via TOPMed. All WHI participants provided informed consent and the study was approved by the IRB of the Fred Hutchinson Cancer Research Center.

The HCHS/SOL was used to establish association chains that include an SDB trait, a transcript, and a metabolite. It is a longitudinal cohort study of U.S. Hispanics/Latinos (30, 31). The HCHS/SOL baseline exam occurred on 2008-2011, where 16,415 participants were enrolled.

HCHS/SOL individuals who consented further participated in an in-home sleep study, using a validated type 3 home sleep apnea test recording airflow (via nasal pressure), oximetry, position, and snoring. Genetic data were measured and imputed to the TOPMed freeze 5b reference panel, as previously described, for individuals who consented at baseline (32, 33). Metabolomic data were also measured for n=∼4,000 individuals selected at random out of those with genetic data (34). **Figure S2** in the Supplementary Information provides the data flow in HCHS/SOL, focusing on individuals with genetic data and wide consent for genetic data sharing. The HCHS/SOL was approved by the institutional review boards (IRBs) at each field center, where all participants gave written informed consent, and by the Non-Biomedical IRB at the University of North Carolina at Chapel Hill, to the HCHS/SOL Data Coordinating Center. All IRBs approving the study are: Non-Biomedical IRB at the University of North Carolina at Chapel Hill. Chapel Hill, NC; Einstein IRB at the Albert Einstein College of Medicine of Yeshiva University. Bronx, NY; IRB at Office for the Protection of Research Subjects (OPRS), University of Illinois at Chicago. Chicago, IL; Human Subject Research Office, University of Miami. Miami, FL; Institutional Review Board of San Diego State University, San Diego, CA.

### RNA sequencing

For both MESA and WHI, RNA-seq was performed via the Trans-Omics in Precision Medicine (TOPMed) program. In MESA, RNA-seq was generated from three blood cell types: peripheral blood mononuclear cells (PBMCs; ∼n=1,200 measured in blood from visits 1 and 5), and specific components: T-cells, and monocytes (referred to as T-cell and Mono, for both: n=416), measured in blood from visit 5. Samples were sequenced at the Broad Institute and at the North West Genomics Center (NWGC). Both centers used harmonized protocols. RNA samples quality was assessed using RNA Integrity Number (RIN, Agilent Bioanalyzer) prior to shipment to sequencing centers. QC was re-performed at sequencing centers by RIN analysis at the NWGC and by RNA Quality Score analysis (RQS, Caliper) at the Broad Institute. A minimum of 250ng RNA sample was required as input for library construction, performed using the Illumina TruSeq^TM^ Stranded mRNA Sample Preparation Kit. RNA was sequenced as 2x101bp paired-end reads on the Illumina HiSeq 4000 according to the manufacturer’s protocols. Target coverage was of ≥40M reads. Comprehensive information about the RNA-seq pipeline used for TOPMed can be found in https://github.com/broadinstitute/gtex-pipeline/blob/master/TOPMed_RNAseq_pipeline.md under MESA RNA-seq pilot commit 725a2bc. Here we used gene-level expected counts quantified using RSEM v1.3.0 (35). RNA sequencing for WHI (whole blood) was performed at the Broad Institute using the unified TOPMed protocols. More information about RNA-seq in WHI is provided in the Supplemental Information.

### Metabolomics data in HCHS/SOL

Metabolomics profiling using fasting blood samples was conducted at Metabolon (Durham, NC) with Discovery HD4 platform in 2017. Serum metabolites were quantified with untargeted, liquid chromatography-mass spectrometry (LC-MS)-based quantification protocol (36, 37). The platform captured a total of 1,136 metabolites, including 782 known and 354 unknown (unidentified) metabolites. Detailed methodologic information is provided elsewhere (34).

### Phenotypic measures of sleep-disordered breathing (SDB)

We used three SDB measures, as measured by overnight sleep studies in MESA and HCHS/SOL (methods above): (1) the Apnea-Hypopnea Index (AHI), defined in MESA as the number of apneas (breathing cessation) and hypopneas (at least 30% reduction of breath volume, accompanied by 3% or higher reduction of oxyhemoglobin saturation) per hour of sleep, and in HCHS/SOL, due to differences in the recording montage compared to MESA, as the number of apnea or hypopnea events with 3% desaturation per hour of sleep; (2) minimum oxyhemoglobin saturation during sleep (MinO2), and (3) average oxyhemoglobin saturation during sleep (AvgO2).

### Testing the association between SDB and blood cell-specific transcriptome-wide gene expression

We used the Olivia R package (38) to perform association analyses of gene expression in PBMCs, monocytes, and T-cells with each of the three SDB measures, separately and in a joint analysis in MESA. SDB phenotypes were treated as the exposures. We followed the recommended Olivia pipeline. Briefly, we performed median normalization, and then filtered lowly expressed gene transcripts defined by removing transcripts with proportion of zero higher than 0.5, median value lower than 1, maximum expression range value lower than 5, and maximum expression value lower than 10. Transcript counts were log transformed after counts of zero were replaced with half the minimum of the observed transcript count in the sample. The analyses were adjusted for age, sex, study center, race/ethnic group, and batch variables: plates, shipment batch, and study site. Because BMI is a strong risk factor for SDB and is assumed to be part of the causal chain, we conducted additional analyses adjusting for BMI (BMIadj). We computed empirical p-values to account for the highly skewed distribution of SDB phenotypes, which may lead to false negative associations if ignored. Finally, we accounted for multiple testing by applying False-Discovery Rate (FDR) correction to each of the association analyses using the Benjamini-Hochberg (BH) procedure (39). We carried forward transcript associations with FDR p-value<0.1 for additional analyses and visualized their association with SDB phenotypes via a hierarchically-clustered heatmap.

### Transcript polygenic risk scores (tPRS) construction and validation

To develop tPRS, we first performed a genome-wide association study (GWAS) for each SDB-associated transcript using the MESA TOPMed WGS dataset; each GWAS adjusted for age (years), sex, study site, self-reported race/ethnic background, and 11 principal components, and analyses were restricted to genetic variants with a minor allele frequency of at least 0.05 (due to low sample size). For each GWAS, we used the fully-adjusted two-stage procedure for rank-normalizing residuals in association analyses (40) to identify genetic variants associated with transcript expression. For PBMCs, we used transcript measures from the two MESA visits with RNA-seq data to increase power. To do this, we removed related individuals, and used a random effect model that accounted for individuals. Summary statistics from the GWAS for each transcript were used to develop PRS weights for the corresponding transcript. Next, we constructed tPRS in MESA. We applied clump and threshold implemented in PRSice2 v2.3.1.e (41) using clumping parameters R^2^=0.1, distance of 250Kb, and three p-value thresholds (5x10^-8^,10^-7^, 10^-6^). For each transcript, we constructed the three tPRS in WHI. A tPRS with the smallest p-value in association with the transcript in WHI, and also having p-value<0.05/3=0.017, was selected and considered validated. We also computed FDR-adjusted p-values based on all constructed tPRS (3 candidate tPRS per gene across all genes). To test the association of the tPRS with transcript in WHI, we used logistic mixed models, executed with the GENESIS R package (42) version 2.16.1. Each tPRS served as the exposure, and transcripts served as the outcome, here too using the two-stage procedure for rank-normalization (40). Relatedness was modeled via a sparse kinship matrix among TOPMed WHI individuals. We selected transcripts with p-value <0.017 for follow-up analysis.

We validated that our approach to construct tPRS is robust. We compared a few polygenic prediction models developed using bulk RNAseq in monocytes. First, the prediction model developed using prediXcan based on the MESA dataset (43, 44), with weights provided in the predictDB database (http://www.predictdb.org/). Second, our approach above using genome-wide SNPs (including trans-eQTLs), and third, a similar clump and threshold approach as above limited to cis-eQTLs defined as SNPs within 1Mbp of the start and end position of the transcript (the definition used by prediXcan). We focused on monocytes for this comparison because prediXcan models were only published based on monocytes.

### Using tPRS to identify reverse association between gene expression and SDB traits

We constructed generalized tPRS in HCHS/SOL. We used HCHS/SOL genotypes imputed to the TOPMed freeze5b reference panel. Prior to tPRS construction, we filtered SNPs with imputation quality <0.8, minor allele frequency <5%, missingness rate >0.01. As illustrated in column B of **Figure 2**, We identified potential reverse causation, where gene expression alters SDB, by using the tPRS constructed in HCHS/SOL as instrumental variables (IVs) and testing their association with their respective SDB phenotypes in HCHS/SOL. We used logistic mixed models, executed with the GENESIS R package (42) version 2.16.1. Each tPRS served as the exposure, and the relevant SDB phenotype served as the outcome. To account for skewness of the SDB phenotypes, we used the two-stage procedure for rank-normalization (40). Relatedness was modeled via a sparse kinship matrix, household sharing, and block unit sharing among HCHS/SOL individuals. Association analyses were adjusted for age, sex, study site, Hispanic/Latino background, the first 5 PCs of the genetic data, and log of the sampling weights used to sample HCHS/SOL individuals into the study. Because the tPRS represent a genetic proxy for gene expression, if a tPRS was found to be associated with a SDB phenotype (p-value<0.05), it provided evidence that the transcript contributed to the SDB phenotype, rather than vice versa. However, as illustrated in diagrams A.2 and A.3 in **Figure 2** for sleep-transcript association, bidirectional associations are also plausible.

### Associations between tPRS and metabolites

Treating tPRS as genetic IVs for gene expression, we estimated associations between tPRS and all identified (named) metabolites with < 25% missing values in HCHS/SOL. We used robust survey models implemented in the R survey package version 4.0 (45), accounting for HCHS/SOL study design (probability sampling and clustering) and providing associations generalizable to the HCHS/SOL target population. For each metabolite, we first imputed observations with missing values of that metabolite with its minimum value observed in the sample, under the assumption that missing values are due to concentrations being below the detection limit, and then rank-normalized it across the sample. We used the same covariates as before: age, sex, study site, Hispanic background, and the first 5 PCs of the genetic data.

Furthermore, we adjusted for BMI depending on the original association of the SDB phenotype and the transcript (BMI unadjusted or BMI adjusted). For each transcript, we corrected metabolite associations to account for FDR using the Benjamini-Hochberg (BH) procedure (39). Associations were considered significant if the FDR p-value was <0.05.

### Association analyses of SDB traits with selected metabolites to verify a complete association chain

To further validate a complete association “chain” as detailed in **Figure 2**, we performed association analyses between the SDB phenotypes and metabolites identified in the tPRS analysis. Associations between SDB phenotypes and metabolites used a survey sampling approach to account for HCHS/SOL sampling design and obtained estimates generalizable to the HCHS/SOL target population. Thus, we used the survey R package (46) with each individual weighted by their sampling weights, and clustering accounted for when computing robust standard errors. Analyses were adjusted for age, sex, study site, Hispanic/Latino background. and BMI depending on the original detected SDB-transcript association (BMI unadjusted or BMI adjusted). If an SDB phenotype was associated with the metabolite (p-value<0.05), we interpreted this as validation of a SDB association with this metabolite via the transcript-level chain.

## RESULTS

### Sample characteristics

Characteristics of the MESA population that participated in the TOPMed omics study, the sleep study, and the smaller T-cells and monocytes analyses are provided in **Table S1**; characteristics of the HCHS/SOL participants with genetic and metabolite data are provided in **Table S2**.

MESA individuals are a multi-ethnic sample, 69 years old on average during MESA exam 5, and 52% female. HCHS/SOL individuals are from diverse Hispanic/Latino backgrounds with a mean age of 46 years during the baseline exam, and 59% female. SDB phenotypes were more severe in MESA, with average AHI=18.6, MinO2=83, and AvgO2=94.1, in contrast to HCHS/SOL with average AHI=6.4, MinO2=87.1, and AvgO2=96.4, consistent with the older age of the MESA sample. Characteristics of the WHI participants with RNA-seq data used to validate the transcript PRS are provided in **Table S3**. WHI individuals are from a multi-race and ethnic sample and are 80 years old on average at the Long-Life Study exam when RNA was extracted, and are all females.

### SDB phenotypes for oxyhemoglobin saturation and AHI are linked to tissue-specific changes in the transcriptome

In MESA, we identified 96 and 24 differentially expressed transcripts (**Table S4, S5** and **Table S6, S7**) with FDR p-value < 0.1 in unadjusted and adjusted BMI analyses, respectively, in the different cell types. **Table 1** reports the top differentially expressed transcripts (FDR p-value <0.05). Three transcripts, *AJUBA* (Ajuba LIM Protein), *ZNF665* (Zinc Finger Protein 665), and *TMC3-AS1* (TMC3 Antisense RNA 1, a long non-coding RNA), are significantly associated with AvgO2 and AHI in both analyses, in the direction of reduced expression with worse SDB measures (higher AHI, lower AvgO2).

Top results from the tissue-specific transcriptome-wide gene expression analysis of SDB phenotypes (FDR p<0.05) in MESA.

**Table 1. Top results from the tissue-specific transcriptome-wide gene expression analysis of SDB phenotypes (FDR $p < 0.05$ ) in MESA.**
| Unadjusted for BMI |  |  |  |  |  |
| --- | --- | --- | --- | --- | --- |
| Gene | Adj LFC | P-value | FDR p-value | SDB Trait | Cell Type |
| AJUBA | 0.104 | 2.59E-06 | 0.050 | AvgO2 | PBMCs |
| PDGFC | -0.016 | 2.59E-06 | 0.050 | MinO2 | PBMCs |
| SIAE | -0.011 | 1.26E-06 | 0.020 | MinO2 | Monocytes |
| EMP1 | -0.030 | 6.91E-06 | 0.050 |  |  |
| LHFPL2 | -0.014 | 9.42E-06 | 0.050 |  |  |
| ZNF665 | -0.011 | 2.74E-07 | 0.003 | AHI | T-cells |
| FAM106A | -0.032 | 7.69E-06 | 0.047 |  |  |
| TMC3-AS1 | -0.022 | 2.74E-07 | 0.003 |  |  |
| Adjusted for BMI |  |  |  |  |  |
| Gene | Adj LFC | P-value | FDR- p-value | SDB Trait | Cell Type |
| AJUBA | 0.116 | 2.59E-07 | 0.005 | AvgO2 | PBMCs |
| ZNF665 | -0.012 | 5.49E-07 | 0.010 | AHI | T-cells |
| DUX4L27 | -0.029 | 6.59E-06 | 0.043 |  |  |
| TMC3-AS1 | -0.021 | 7.14E-06 | 0.043 |  |  |
| The table provides results from analyses of unadjusted (n=8 transcripts) and adjusted (n=4 transcripts) for BMI. “Adj LFC” is the covariate-adjusted log2-fold change in gene expression per 1 unit increase in SDB exposure. P-value is the raw p-value, and FDR p-value is the p-value following FDR adjustment using the Benjamini-Hochberg procedure. For AvgO2 and MinO2, negative Adj LFC indicates increased expression with worse SDB symptoms. For AHI, positive Adj LFC indicates increased expression with worse SDB symptoms. |  |  |  |  |  |

To visualize gene expression and compare across SDB traits and cell types, Spearman correlation of log-fold change in expression of all SDB-associated transcripts (n=96 transcripts FDR p<0.1) was illustrated with a heatmap for BMI unadjusted (**Figure S3**) and BMI adjusted (**Figure S4**) analyses, clustered using hierarchical clustering based on the correlation between the log-fold estimates. These results illustrate concordant and discordant patterns of differential gene expression by cell type (PBMCs, monocytes, and T-cells) and SDB trait (AvgO2, MinO2, and AHI). There are a few striking differences in gene expression, particularly the increased expression of *FAM106A, DNAJA3*, *BCDIN3D-AS1, TGFBRAP1, BEND5, TTC24*, *TMC3-AS1*,

*LINC00235*, *SEC14L2*, *ARHGEF9, TSHZ1*, and *LA16c-312E8.4* in T-cells compared to monocytes and PBMCs. To further investigate the overall patterns in gene expression in relation to tissue type and SDB traits, a heatmap of the Spearman correlation of the log-fold expression estimates of SDB phenotypes was plotted **Figure 3**. Within cell types, the SDB traits AHI and MinO2 had the highest correlation for gene expression (Spearman R^2^ between 0.91 to 0.97), whereas AvgO2 had lower correlations with AHI and MinO2, especially in monocytes. In addition, a heatmap of estimated log-fold gene expression change (FDR p-value < 0.1) with SDB phenotypes across tissues adjusted for BMI is shown in **Figure S5.** The correlations between the SDB effect estimates for gene expression across cell types are different, and generally higher, from the phenotypic correlations between the SDB phenotypes, which are at the range of 0.53 to 0.73 Spearman R^2^ (**Figure S6**). When computing correlations over all genes, estimated associations between AvgO2 with gene expression had almost no correlation with the other phenotypes (**Figure S7**). The same patterns (although slightly attenuated) are observed in BMI adjusted analyses (**Figure S8**).

**Figure 3:**
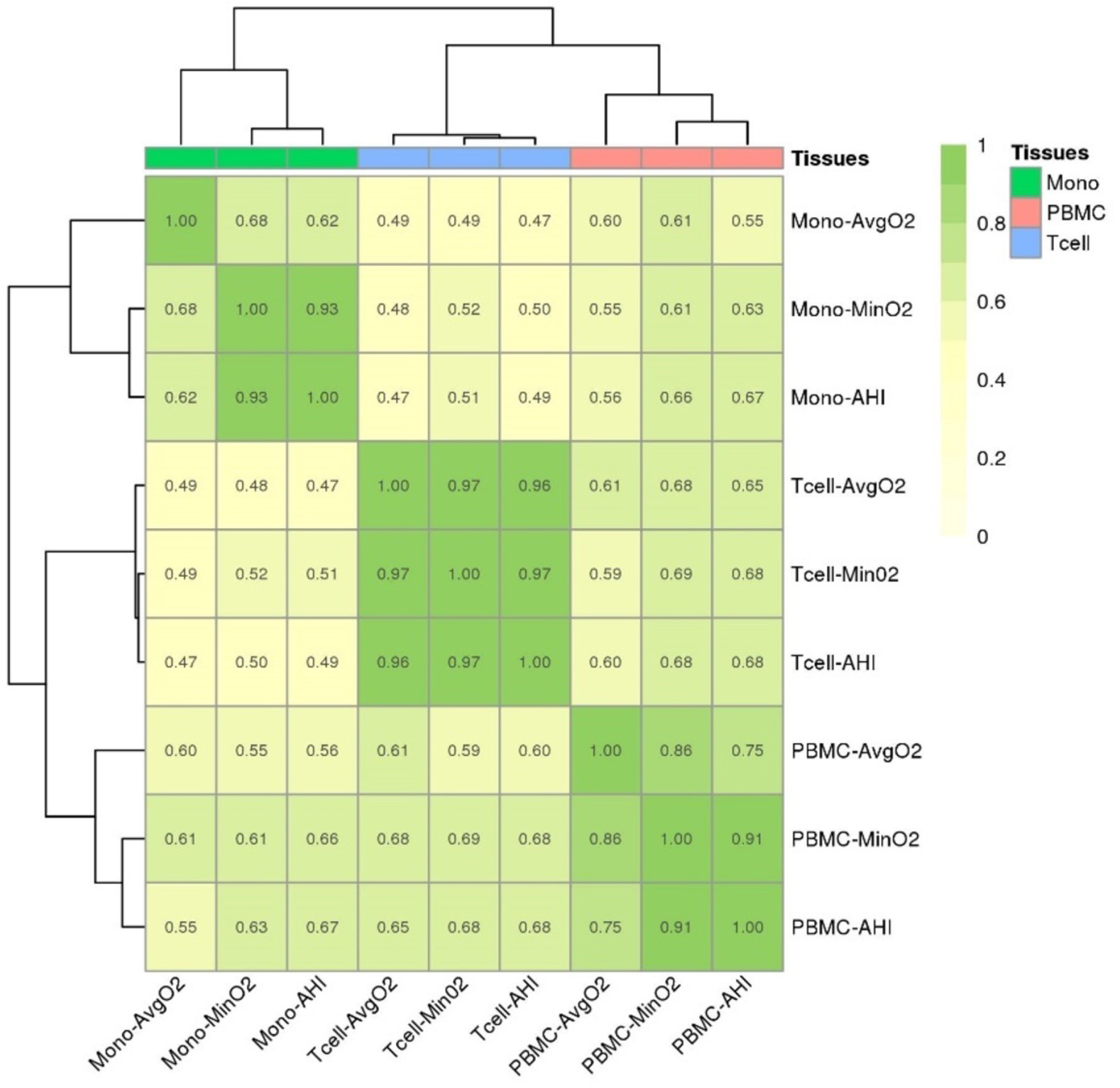
Spearman correlations between estimated log-fold changes in gene expression across SDB phenotypes and blood cell types without BMI adjustment in MESA. Heatmap illustrating the Spearman correlations of log-fold change of transcript expression by tissue type (monocytes, T-cells, PBMCs) and SDB phenotype (AvgO2, MinO2, AHI) in MESA. Correlations were computed over genes with FDR p<0.1. Color legend portrays Spearman R^2^ (no/weak correlation = light yellow; complete/strong correlation = green). Estimated AHI effect sizes were flipped prior to computation of correlations so that they match the direction of MinO2 and AvgO2.

### Construction and validation of transcript PRS

We constructed tPRS for gene expression in monocytes using a few methods, focusing on transcripts that were associated with SDB exposures in our analysis. The performance of constructed tPRS was evaluated against whole-blood gene expression levels in n=1,269 WHI participants. **Figure S9** visualizes the results, demonstrating that tPRS constructed using the clump and threshold for genome-wide SNPs, including trans-eQTLs and tPRS focusing on cis-eQTLs have similar results, and the same generalization rate as that of the prediXcan-based tPRS. However, prediXcan tPRS had opposite direction of association with one of the transcripts in WHI, and, both cis-eQTLs based tPRS (prediXcan and clump and threshold) were not available for some transcripts due to lack of transcript-associated SNPs near the coding region. Thus, we moved forward with the genome-wide approach. Of the 96 tPRS (BMI unadjusted analysis) and 24 tPRS (BMI-adjusted analysis) tested, 26 and 9 tPRS were associated (p<0.017) with gene expression (**Tables S8-S9**) in whole blood and considered validated as IVs.

### Evidence of causal association between transcripts and SDB phenotypes

We tested the association of the validated tPRS, constructed in a cell-specific manner, with SDB phenotypes in HCHS/SOL (**Table S10**). Of the 26 tested in BMI unadjusted analysis, 3 tPRS showed evidence of reverse association with SDB phenotypes (p-value<0.05), supporting a causal relationship between expression of these transcripts and SDB traits. Among them, the strongest association was of the tPRS for *P2RX4* (Purinergic Receptor P2X 4) in PBMC, in its association with AvgO2, one standard deviation (SD) increase in the PRS was associated with increased 1.9% AvgO2. Additionally, tPRS for *SEC14L2* (SEC14 Like Lipid Binding 2) was negatively associated with AHI in T-cells and tPRS for *TUBB6* (Tubulin Beta 6 Class V) was positively associated with MinO2 in monocytes. After BMI adjustment, only *P2RX4* remained positively associated with AvgO2 in PBMCs (p-value <0.05), as shown in **Table S11**.

### Evidence of causal association between transcripts and metabolites

We tested the relation between each validated tPRS and metabolites in HCHS/SOL. The tPRS for P2RX4 and CTD-2366F13.1 (also known as MOCS2-DT, MOCS2 Divergent Transcript) were associated with a total of 6 and 7 metabolites in unadjusted BMI and adjusted BMI analyses (FDR p-value <0.05, Table S12 and Tables S13), respectively; the association “chains” are visualized in **Figure 4**. Of 7 metabolites, 3 of them (butyrylcarnitine, linoleoyl-arachidonoyl-glycerol (18:2/20:4), and palmitoleoyl-linoleoyl-glycerol (16:1/18:2)) were also associated with AvgO2 (Table S15). However, the AvgO2-metabolite associations did not remain after BMI adjustment, suggesting that BMI, rather than SDB, may be driving these associations (**Table S16**). Of the transcripts, P2RX4 had evidence of a complete chain of association with SDB and metabolites (p-value <0.05) in the BMI unadjusted analysis.

**Figure 4.**
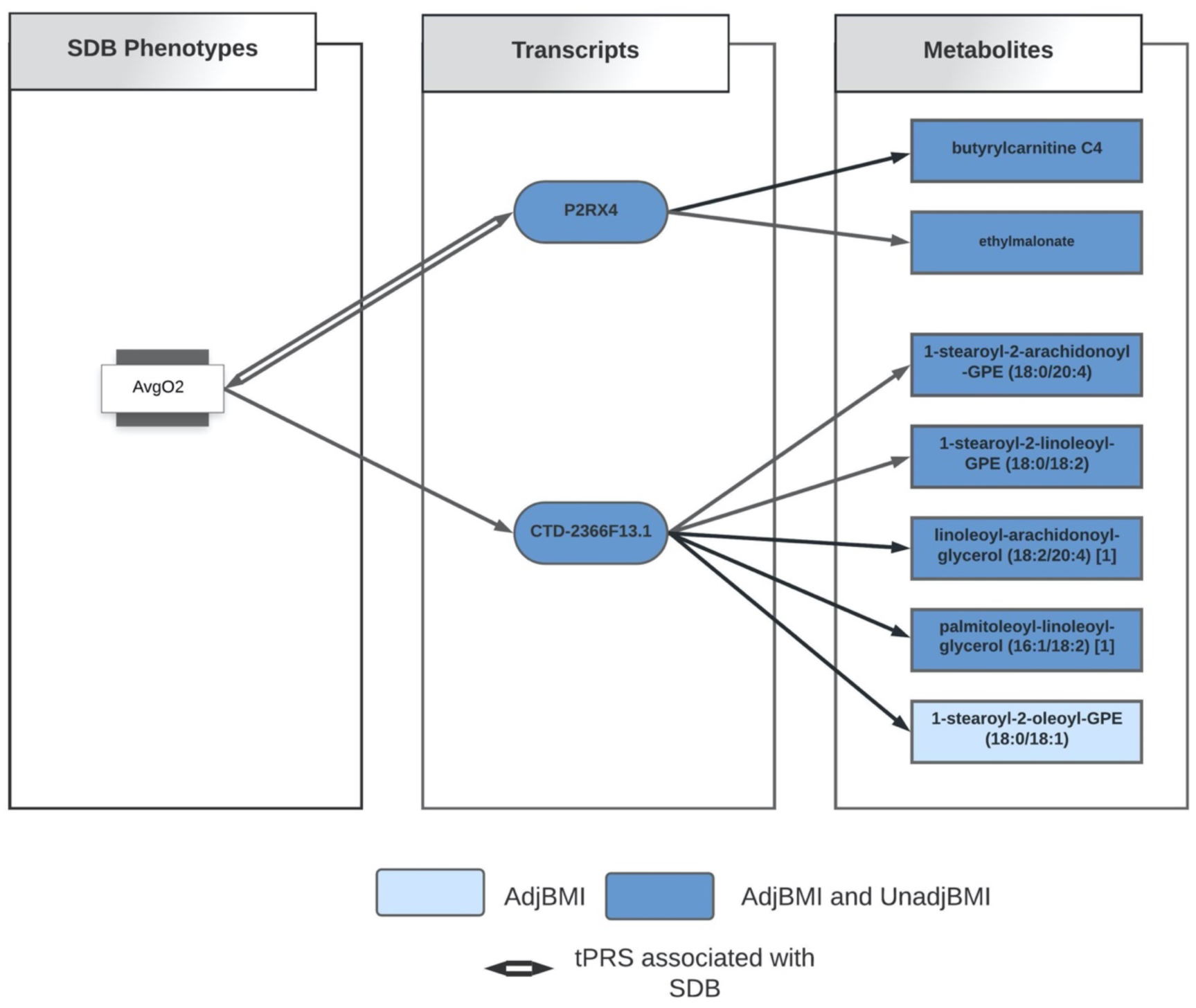
Identified association chains between AvgO2, transcripts, and metabolites. Diagram illustrating the “association chain” relationships between AvgO2, tPRS, and metabolites in BMI unadjusted and BMI adjusted analyses. Dark blue color indicates association between tPRS and metabolites in both BMI unadjusted and adjusted analyses; The light blue color indicates the association between tPRS and metabolite only in BMI adjusted analysis.

## DISCUSSION

Here, we conducted a robust analysis of SDB phenotypes and their multi-omics correlates. We first identified transcriptome-wide tissue-specific changes in gene expression associated with sleep-related oxyhemoglobin saturation traits and AHI in MESA and then used those transcripts to develop genetic proxies for gene expression (tPRS). Next, we generalized and validated some of the tPRS in WHI. Finally, we utilized the validated tPRS to further study SDB phenotypes and metabolite associations in HCHS/SOL. Our results support SDB-related leukocyte alterations in gene expression and highlight signaling pathways related to inflammation, thrombosis, and neurotransmission.

SDB traits were associated with differential expression of many transcripts across three blood cell types (**Table S4 and S5**, 96 genes with FDR p-value<0.1). Of the top transcripts (8 genes with FDR p-value <0.05), *AJUBA* expression was positively associated with higher AvgO2 and *PDGFC* expression was negatively associated with higher MinO2 in PBMCs. *AJUBA* is a scaffold protein in the family of LIM domain-containing protein, considered key regulators of the hypoxic response (47). Recent research supports a role for AJUBA in interacting with retinoic acid receptor signaling in an in vitro model (48) and a role for indirectly limiting inflammation by maintaining mitochondrial quality control in a mouse model (49); therefore, greater *AJUBA* expression may be associated with increased AvgO2 through pathways related to inflammation and retinoic acid. *PDGFC* encodes platelet derived growth factor C (50) and is upregulated during hypoxia in tumor cells (51). The association between higher *PDGFC* expression and lower minimum oxygen saturation (MinO2) supports a role for *PDGFC* signaling in SDB-related hypoxia. In monocytes, *EMP1* was linked to MinO2, and prior studies have shown that *EMP1* expression increases during sleep loss and during hypoxia in cancer tissues (52, 53).

Correlations between leukocyte subsets and trait-specific gene expression (**Figure 3**) supported an overall pattern of similarity between cell populations, but also highlighted some striking differences. For example, the gene *HPCAL4* (Hippocalcin-Like Protein 4) has high expression changes with “worse” SDB phenotypes (higher AHI, lower AvgO2 and MinO2) in monocytes, but weak associations in T-cells and PBMCs. However, *FAM106A* and *SERPINE2* have lower expression with worse SDB phenotypes in T-cells, but weak associations in PBMCs and monocytes. *SERPINE2* (54) encodes glia-derived nexin (GDN, also referred to as protease nexin-1) with mixed evidence as a genetic factor for COPD (55, 56). *SERPINE2/GDN* inhibits the hypoxia-triggered serine protease thrombin (57, 58), and a mouse model of *SERPINE2* deficiency causes excess thrombin activity and overproduction of cytokines in the lungs (59), suggesting a role for *SERPINE2* in airway inflammation. Since OSA may be associated with a procoagulant state featuring high thrombin levels (60, 61), the hypoxemia associated with OSA may lead to increased thrombin levels, affecting *SERPINE2* expression in T-cells.

Of the top differentially expressed genes in MESA whose tPRS was validated in an independent cohort (WHI), only *P2RX4* was found to have a complete “chain” of association with SDB and metabolites when tested in another independent study, HCHS/SOL. The PRS for *P2RX4* was positively associated with AvgO2, both with and without adjustment for BMI, and as such it is a candidate contributor to oxyhemoglobin saturation in SDB. *P2RX4* encodes a purinergic receptor for ATP, P2X4, which may play a role in the neuroprotective effects of hypoxic preconditioning (62, 63). *P2RX4* was further negatively associated with metabolite butyrylcarnitine, an indicator of fatty acid metabolism previously linked to BMI (64). Higher oxygen tension promotes increased ATP production (65), which may in turn promote increased *P2RX4* expression (P2X4 as a purinergic ATP receptor) and decreased butyrylcarnitine (66).

*P2RX4* can have beneficial or detrimental effects depending on context. A mouse model of genetically increased *P2XR4* expression led to enhanced cardiovascular function (67) and prior research supports a role for *P2XR4* in heart contractility (68), suggesting that *P2XR4* may positively drive AvgO2 via bolstering cardiac force. Likewise, butyrylcarnitine was found to be associated with time of cardiac isovolumetric relaxation and may be a marker of heart failure (69), further linking *P2XR4* to cardiac function. While *CTD-2366F13.1* (*MOCS2-DT*) was not associated with SDB traits, two of the four metabolites negatively associated with *CTD-2366F13.1* were also associated with AvgO2: linoleoyl-arachidonoyl-glycerol (18:2/20:4) and palmitoleoyl-linoleoyl-glycerol (16:1/18:2). Levels of 2-Arachidonoylglycerol, an agonist of the CB1 and CB2 cannabinoid receptors, are increased in the brain during ischemia (70) and in macrophages in response to oxidative stress (71). Therefore, greater AvgO2 levels may result in decreased linoleoyl-arachidonoyl-glycerol (18:2/20:4) levels.

There are several strengths and some limitations of our analysis. Our unique study design exploited a stepwise discovery/validation approach across multiple studies and optimized the availability of SDB-related datasets to study omics markers and SDB. First, we identified SDB-related transcripts. Next, we utilized genetic associations with gene expression to construct tPRS, serving as “genetic IVs”: exposure variables that are likely associated with the gene transcripts and are specific to them, thus allowing for downstream association analysis and causal inference using these IVs instead of the transcript themselves (72). The idea of using genetic variants as IVs is often used in Mendelian Randomization (MR) analysis. Our analysis is different than standard one-sample MR in that we did not estimate the effect of the transcript on the outcome, because we did not have access to RNA-seq in HCHS/SOL. However, for causality inference, it is sufficient to test the IV association with the outcome of interest (73). We then studied the evidence for the effect of gene expression on SDB using the tPRS. Still, the exact form of association between the gene expression and SDB traits cannot be determined (**Figure 2**). For example, if no tPRS-SDB association was detected, it is possible that this was due to lack of power. Even in the absence of tPRS-SDB association, the association between the SDB and tPRS can be due to either causal effect of SDB on tPRS, or confounding by a common cause of both. Notably, during the WHI validation step, many transcripts did not have significant tPRS associations and therefore were not carried forward for the genetic association analysis in HCHS/SOL; lack of validation may be due to cell type differences, as we validated tPRS in WHI, where gene expression was measured in whole blood, unlike measurement in specific cell types in MESA. Finally, we leveraged the validated tPRS to test for associations of gene transcript expression with metabolites and connect possible “chains” of associations. All included cohorts are large and represent diverse populations in the U.S. Our sleep cohorts, HCHS/SOL and MESA, have objective sleep phenotype measurement without prior selection of participants based on specific phenotypes. Other limitations of our study include high multiple testing burden, performing procedures with multiple steps, utilizing multiple data in constructing tPRS, and differences in sample timing between blood sample collection and overnight PSG in MESA. However, genetic data should not be affected by differences in timing, and chronic conditions like SDB may be stable over time, making this limitation less of a concern. It is notable that the three blood cell types used in MESA are not distinct: PBMCs include monocytes and T-cells. Further, both monocytes and T-cells are also composed of more granular cell types. Statistical analyses within one cell type are generally powered to detect associations that hold across the component, more granular, cell types, and some cell type-specific associations may be masked. Overall, we utilize robust statistical methods and objective measures, integrating across multiple layers of biological measures, to interrogate the mechanisms driving SDB-related morbidity.

## CONCLUSION

In summary, we examined multiple levels of biological information to investigate signaling mechanisms underlying SDB traits to better understand drivers of morbidity in SDB. Our results highlight differential gene expression by circulating leukocyte populations in relation to multiple SDB traits related to hypoxia, neurotransmission, and thrombolytic activity. Analyses with validated tPRS in independent cohorts support a mechanistic role for *P2XR4* purinergic signaling in SDB, a gene known to influence cardiac function, which is relevant to SDB as both a risk factor and outcome. Overall, we applied novel, robust methods to integrate multi-omic data and SDB data to discover mechanisms underlying multiple SDB traits. Our multi-dimensional approach using large population cohorts is a promising approach to unravel biological underpinnings of complex human disorders.

## Data Availability

DATA AVAILABILITY STATEMENT 
MESA, HCHS/SOL and WHI data are available through application to dbGaP according to the study specific accessions. MESA phenotypes are available in: phs000209; WHI phenotypes: phs000200; and HCHS/SOL phenotypes: phs000810. HCHS/SOL genotyping data: phs000880. MESA and WHI RNA-seq data has been deposited and will become available through the TOP-Med according to the study specific accessions; MESA: phs001416 ; WHI:  phs000972. HCHS/SOL metabolomics data are available via data use agreement with the HCHS/SOL Data Coordinating Center at the University of North Carolina at Chapel Hill, see collaborators website: https://sites.cscc.unc.edu/hchs/. Data needed to construct the tPRS are publicly available on the repository  https://github.com/nkurniansyah/SDB_Multi_Omics.

## ACKNOWLEDGEMENTS

The authors thank the staff and participants of HCHS/SOL, MESA, and WHI, for their important contributions. We gratefully acknowledge the investigators and participants who provided biological samples and data for TOPMed.

## AUTHOR CONTRIBUTIONS

T.S. supervised the research. N.K. and T.S. conceptualized and conducted the biostatistical and bioinformatics analyses. N.K., T.S., and D.A.W. interpreted the results and wrote and edited the manuscript. Y.Z., B.Y, B.C., H.W., H.M.O-B, A.P.R., A.R.R., J.D.S., J.C., M.D., P.C.Z., R.K., C.K., S.S.R., J.I.R., S.A.G, and S.R. read and approved the final manuscript.

## DATA AVAILABILITY STATEMENT

MESA, HCHS/SOL and WHI data are available through application to dbGaP according to the study specific accessions. MESA phenotypes are available in: phs000209; WHI phenotypes: phs000200; and HCHS/SOL phenotypes: phs000810. HCHS/SOL genotyping data: phs000880. MESA and WHI RNA-seq data has been deposited and will become available through the TOP-Med according to the study specific accessions; MESA: phs001416 ; WHI: phs000972.

HCHS/SOL metabolomics data are available via data use agreement with the HCHS/SOL Data Coordinating Center at the University of North Carolina at Chapel Hill, see collaborators website: https://sites.cscc.unc.edu/hchs/. Data needed to construct the tPRS are publicly available on the repository https://github.com/nkurniansyah/SDB_Multi_Omics.

## Supplemental Methods

### The Multi-Ethnic Study of Atherosclerosis (MESA)

MESA is a longitudinal cohort study (27), established in 2000, that prospectively collected risk factors for development of subclinical and clinical cardiovascular disease among participants in six field centers across the United States (Baltimore City and Baltimore County, MD; Chicago, IL; Forsyth County, NC; Los Angeles County, CA; Northern Manhattan and the Bronx, NY; and St. Paul, MN). The 1^st^ and 5^th^ MESA exams took place between 2000-2002, and 2010-2012, respectively, and whole blood was drawn from participants in both exams. For about 1,400 participants, blood was used later for RNA extraction and/or proteomics in at least one of the exams. In addition, a sleep study ancillary to MESA occurred shortly after MESA exam 5 during 2010-2013. Sleep study participants underwent single night in-home polysomnography (Compumedics Somte Systems, Abbotsville, Australia, AU), as previously described (28). The number of individuals with each type of data and at each time point (exam 1 and exam 5) varies. **Figure S1** in the Supplementary Information visualizes the data flow and overlaps across the various measures used in this study: whole-genome genotyping, RNA-seq, and sleep. The study was approved by Institutional Review Boards in all study centers and participants provided written informed consent.

### The Hispanic Community Health Study/Study of Latinos (HCHS/SOL)

The HCHS/SOL is a longitudinal cohort study of U.S. Hispanics/Latinos (30, 31) recruited from four geographic regions: Bronx NY, Chicago IL, Miami FL, and San Diego CA. The HCHS/SOL baseline exam occurred on 2008-2011, where 16,415 participants were enrolled via multi-stage probability sampling. HCHS/SOL individuals who consented further participated in an in-home sleep study, using a validated type 3 home sleep apnea test recording airflow (via nasal pressure), oximetry, position, and snoring (ARES Unicorder 5.2; B-Alert). Genetic data were measured and imputed to the TOPMed freeze 5b reference panel as previously described, for individuals who consented at baseline (32, 33). More information about genotyping and imputation is provided in the Supplementary Information. Metabolomic data were also measured for n=∼4,000 individuals selected at random out of those with genetic data (34). **Figure S2** in the Supplementary Information provides the data flow in HCHS/SOL, focusing on individuals with genetic data and wide consent for genetic data sharing. All participants provided written informed consent at their recruitment site and the study was approved by the institutional review boards at all participating institutions.

### Genotyping and imputation in HCHS/SOL

Blood was drawn from HCHS/SOL participants during the baseline exam. Individuals who consented to genetic studies were genotyped using an Illumina Omni2.5M array, which included 150,000 custom-selected Single Nucleotide Polymorphisms (SNPs) including ancestry-informative and Amerindian-specific variants. Global ancestry proportions measuring the proportion of the genome inherited from European, African, and Amerindian ancestors and genetic principal components were computed as previously reported [1]. The genotypes were imputed to the Trans-Omics in Precision Medicine (TOPMed) freeze 5b reference panel as described in [2].

### The Women’s Health Initiative (WHI)

The WHI is a prospective national health study focused on identifying optimal strategies for preventing chronic diseases that are the major causes of death and disability in postmenopausal women [3]. The WHI initially recruited 161,808 women between 1993 and 1997 with the goal of including a socio-demographically diverse population with racial/ethnic minority groups proportionate to the total minority population of US women aged 50-79 years. The WHI consists of two major parts: a set of randomized Clinical Trials and an Observational Study. The WHI Clinical Trials (CT; N=68,132) includes three overlapping components, each a randomized controlled comparison: the Hormone Therapy Trials (HT), Dietary Modification Trial, and Calcium and Vitamin D Trial. A parallel prospective observational study (OS; N = 93,676) examined biomarkers and risk factors associated with various chronic diseases. While the HT trials ended in the mid-2000s, active follow up of the WHI-CT and WHI-OS cohorts has continued for over 25 years with the accumulation of large numbers of diverse clinical outcomes, risk factor measurements, medication use, and many other types of data.

A total of 11,071 WHI participants have whole-genome sequencing data via TOPMed, and 1,274 of these participants have RNA-seq measured in venous blood via TOPMed.

### RNA sequencing in WHI

RNA-seq was performed via the Trans-Omics in Precision Medicine (TOPMed) program. The WHI RNA samples (N=1,335) were collected from Long Life Study (LLS) participants as part of the LLS Blood Protocol using the PreAnalytiX PAXgene blood tubes, a collection system designed to preserve RNA from whole blood. After collection in participant’s homes throughout the US, PAXgene tubes were the last of five tubes drawn from each participant, mixed carefully (inverted 8-10 times), kept at room temperature for a minimum of 2 hours post draw, and shipped overnight with cool packs to the Fred Hutch Specimen Processing Lab (SPL). Upon receipt at the SPL, PAXgene tubes were stored at -80 degrees C until they could be transferred to the Fred Hutch Public Health Sciences Biomarker Lab, where the vials were kept frozen at -80 degrees C. Within about a month of collection, the lab extracted total RNA, including miRNA, using the PreAnalytiX method (*PAXgene Blood miRNA Kit Handbook, Qiagen, 05/2009)* designed for use with the PAXgene blood collection tubes. A qualitative assessment by agarose gel electrophoresis of RNA integrity was done at the time of extraction. The RNA was quantified by NanoDrop. The elution volume of 76 μL of extracted RNA was divided between two RNA ‘Parent’ vials without further dilution, frozen at -80 degrees C, and shipped overnight on dry ice to the WHI biorepository for long-term storage at -80 degrees C. RNA sequencing for WHI was performed at the Broad Institute using the unified TOPMed protocols. More information about RNA sequencing protocols in TOPMed is available here https://github.com/broadinstitute/gtex-pipeline/blob/master/TOPMed_RNAseq_pipeline.md.

---

**Figure S1:**
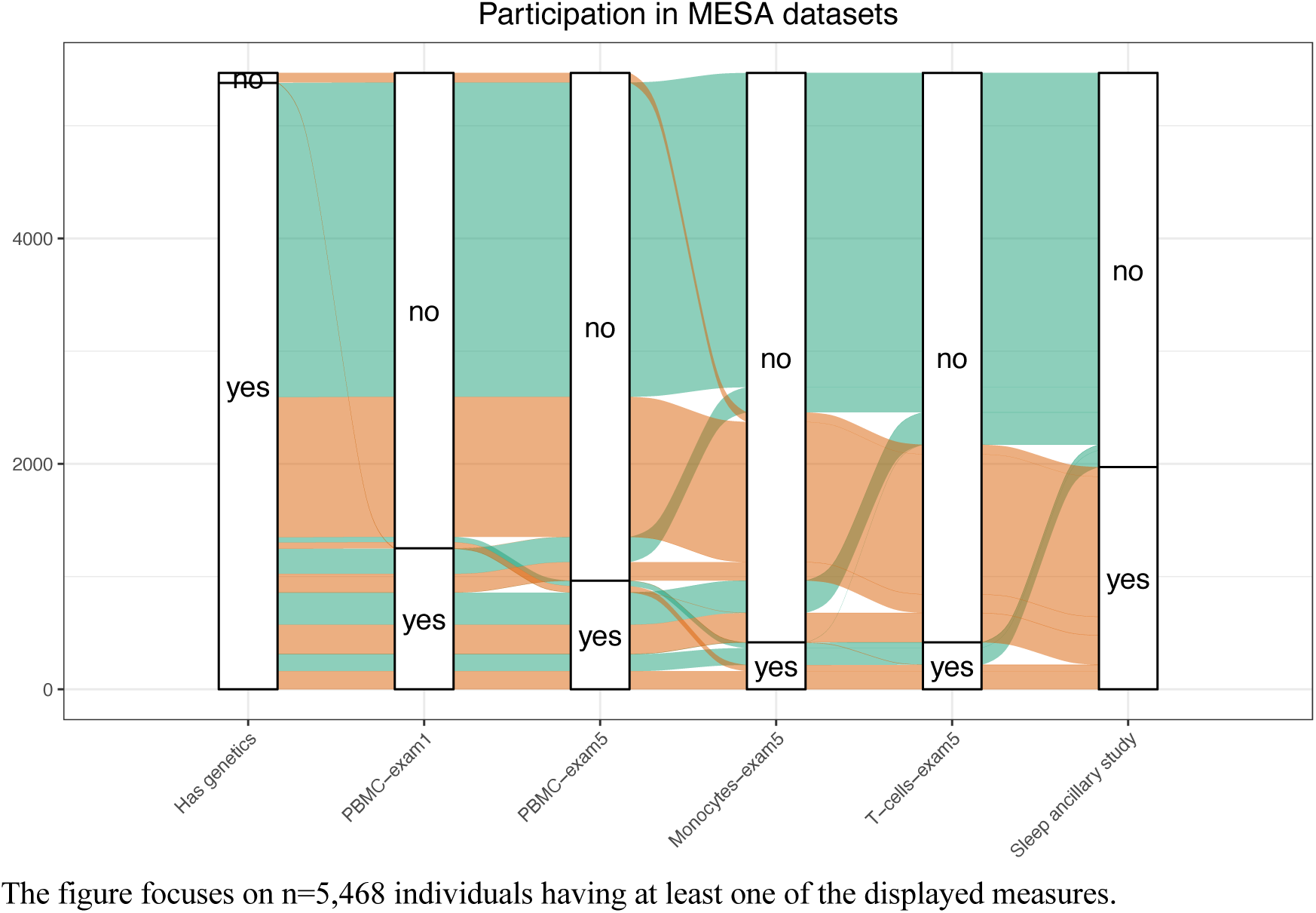
MESA data flow across various measures in the 1^st^ and 5^th^ exam, and the sleep ancillary study.

**Figure S2:**
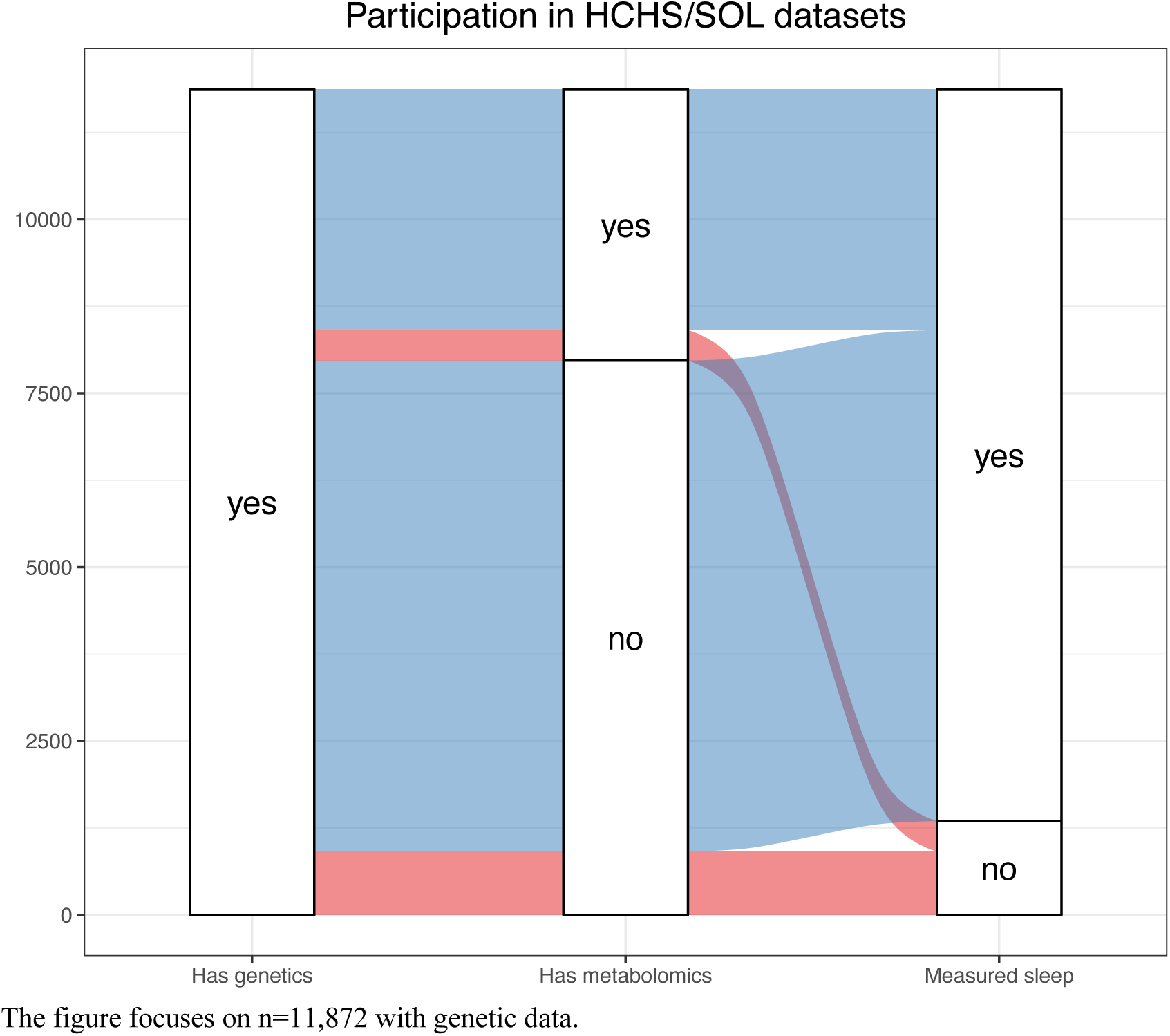
HCHS/SOL data flow across genotyping, metabolomics, and sleep data.

**Figure S3:**
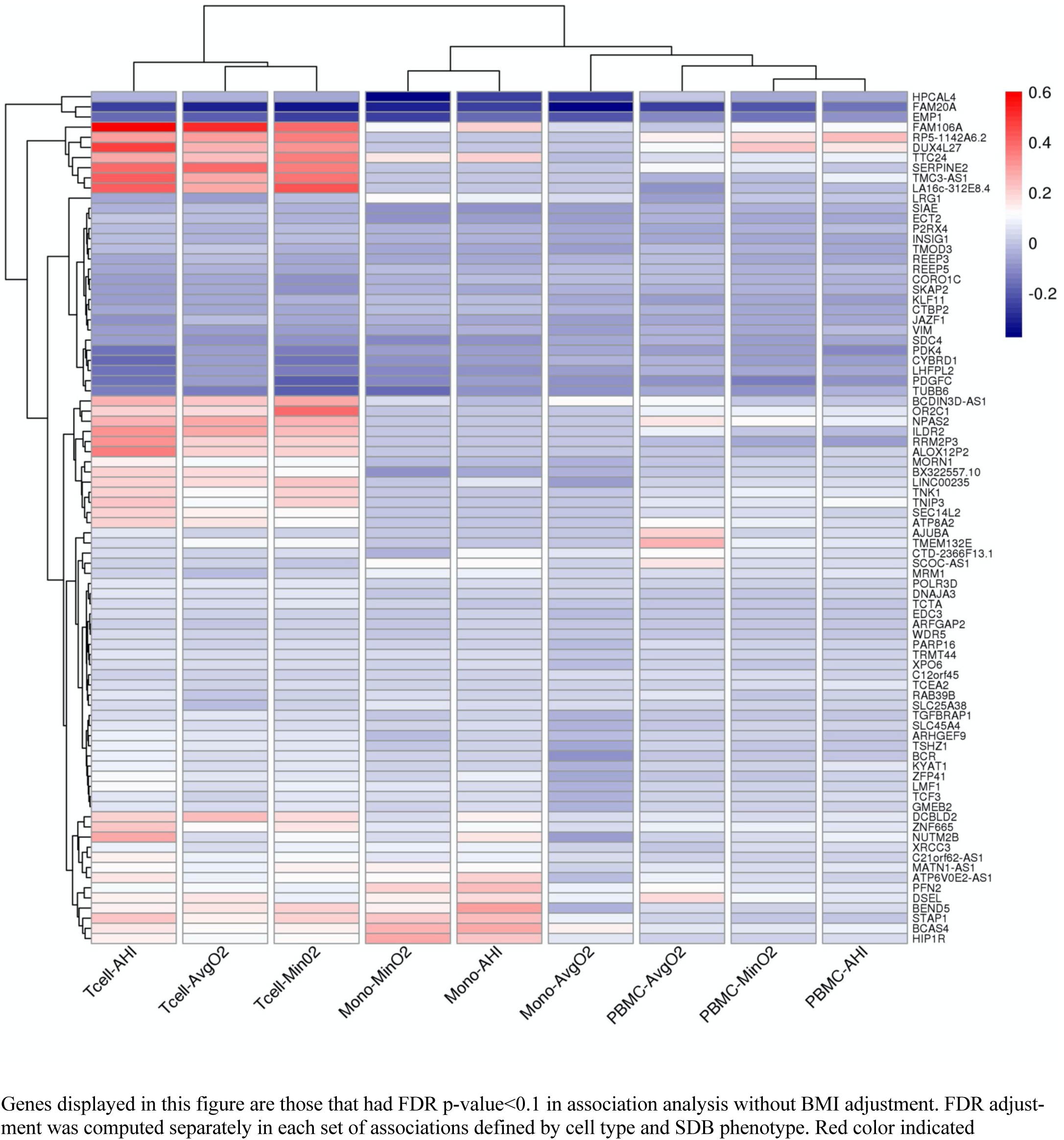
Heatmap of estimated log-fold gene expression change with SDB phenotypes across tissues without BMI adjustment in MESA.

**Figure S4:**
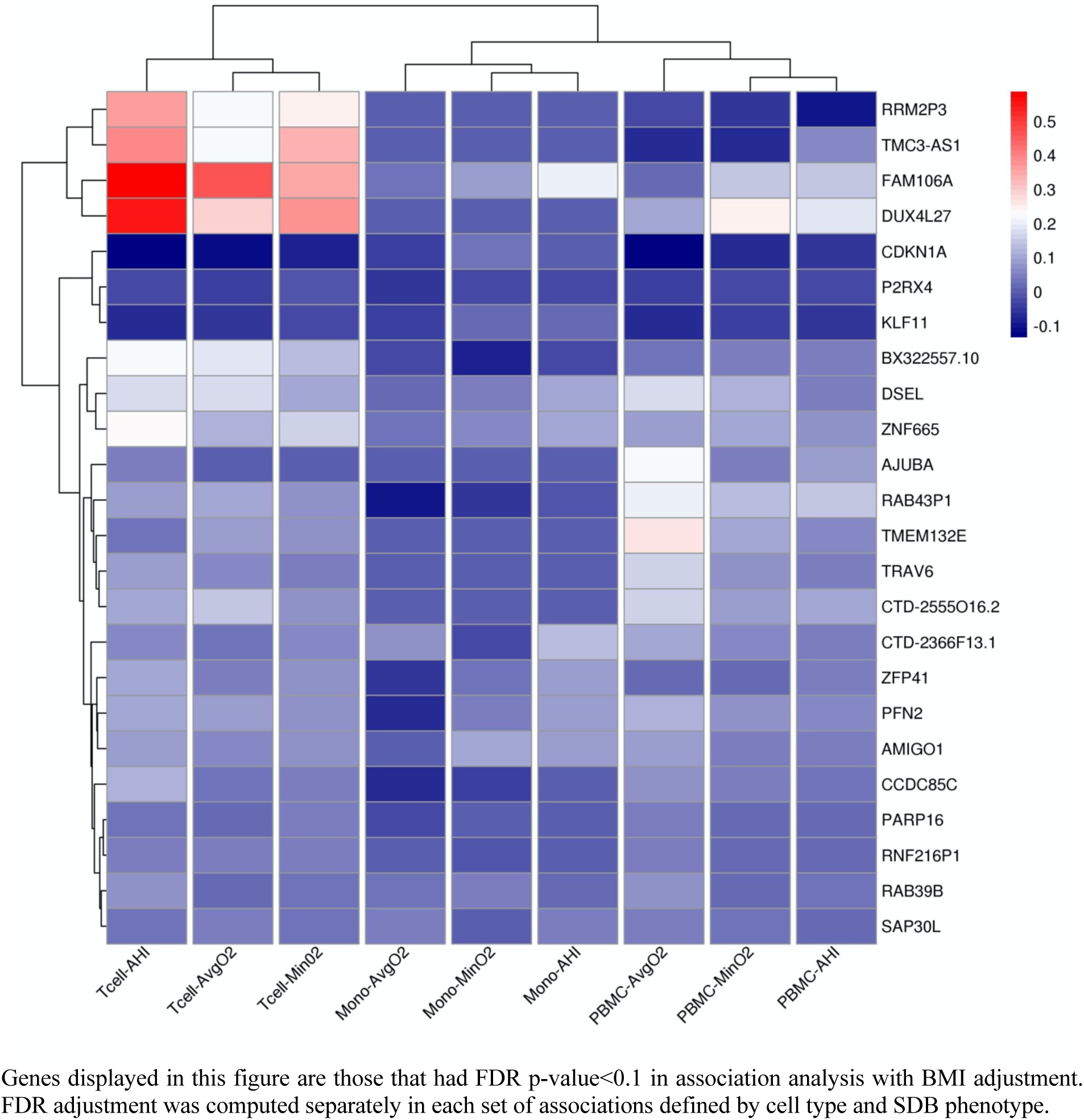
Heatmap of estimated log-fold gene expression change with SDB phenotypes across tissues from BMI-adjusted analysis in MESA.

**Figure S5:**
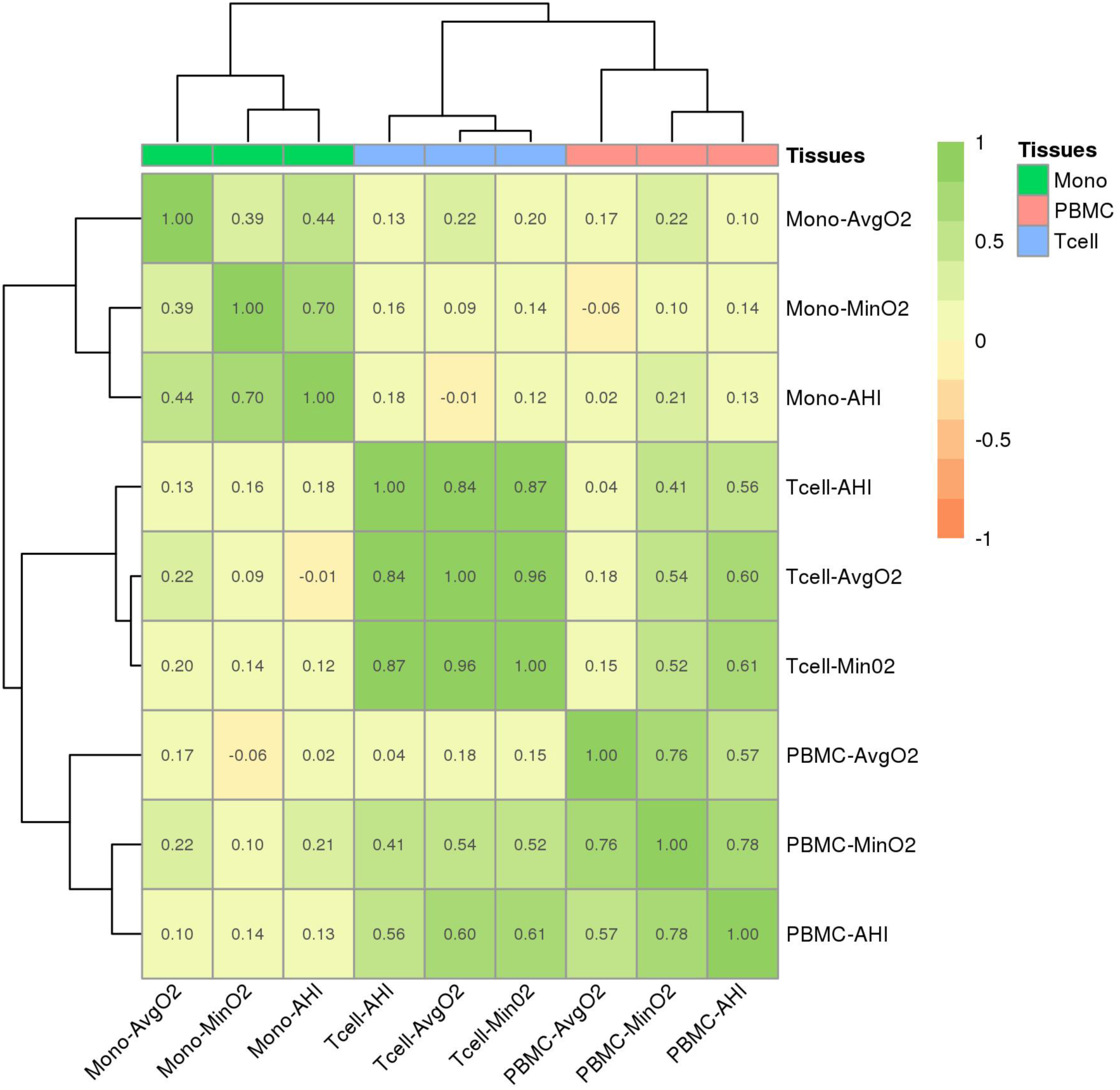
Spearman correlations between estimated log-fold changes in gene expression across SDB phenotypes and tissues in BMI adjusted analysis in MESA (top genes) Heatmap illustrating the Spearman correlations of log-fold change of transcript expression by tissue type (monocytes, T-cells, PBMCs) and SDB phenotype (AvgO2, MinO2, AHI) in MESA. Correlations were computed over genes with FDR p<0.1. Color legend portrays Spearman R^2^ (no/weak correlation = light yellow; complete/strong correlation = green). Estimated AHI effect sizes are flipped prior to computation of correlations so that they match the direction of MinO2 and AvgO2.

**Figure S6.**
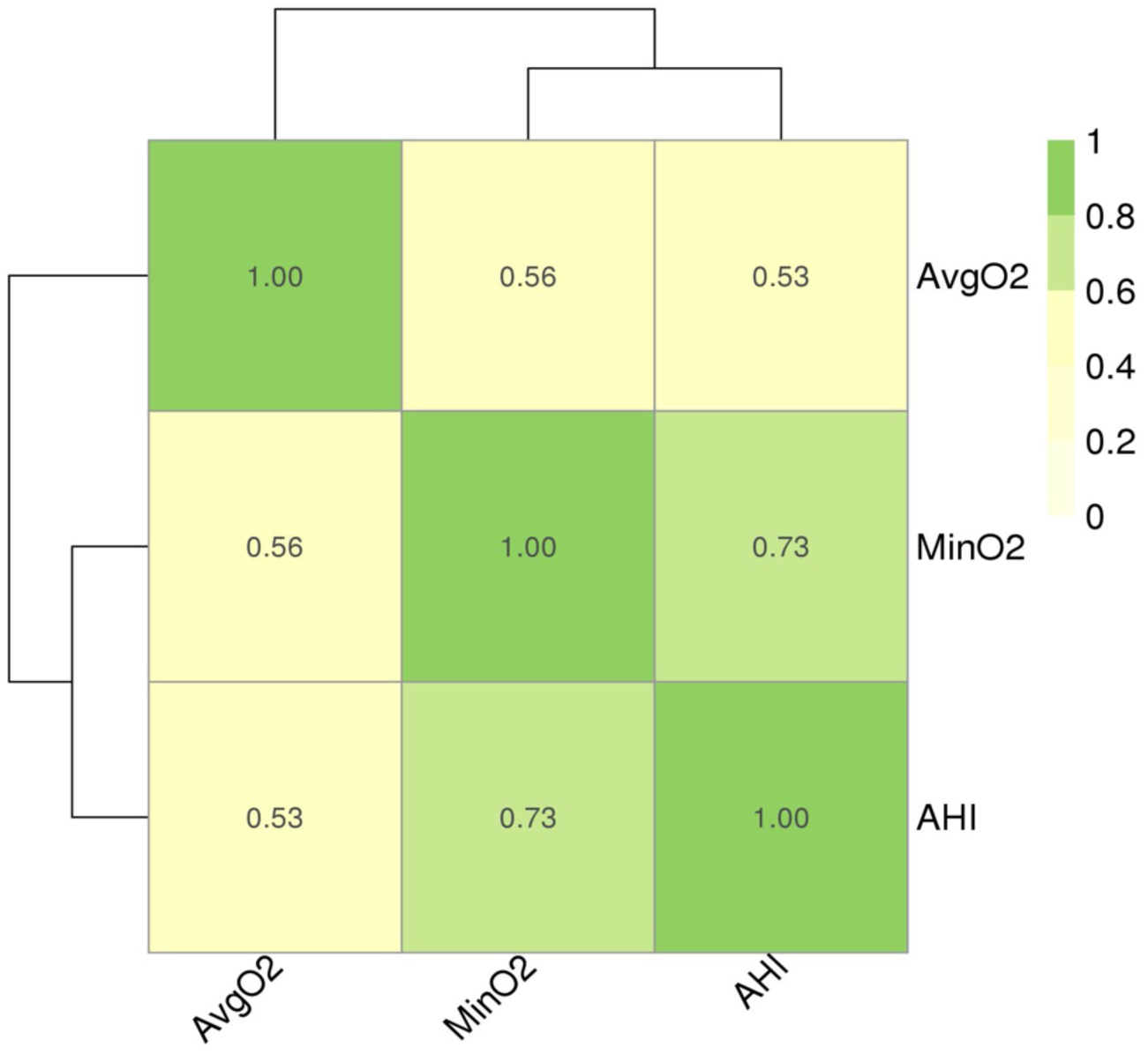
Correlation between the SDB phenotypes. Heatmap illustrating the Spearman correlations SDB phenotype (AvgO2, MinO2, AHI). Color legend portrays Spearman R^2^ (no/weak correlation = light yellow; complete/strong correlation = green).

**Figure S7:**
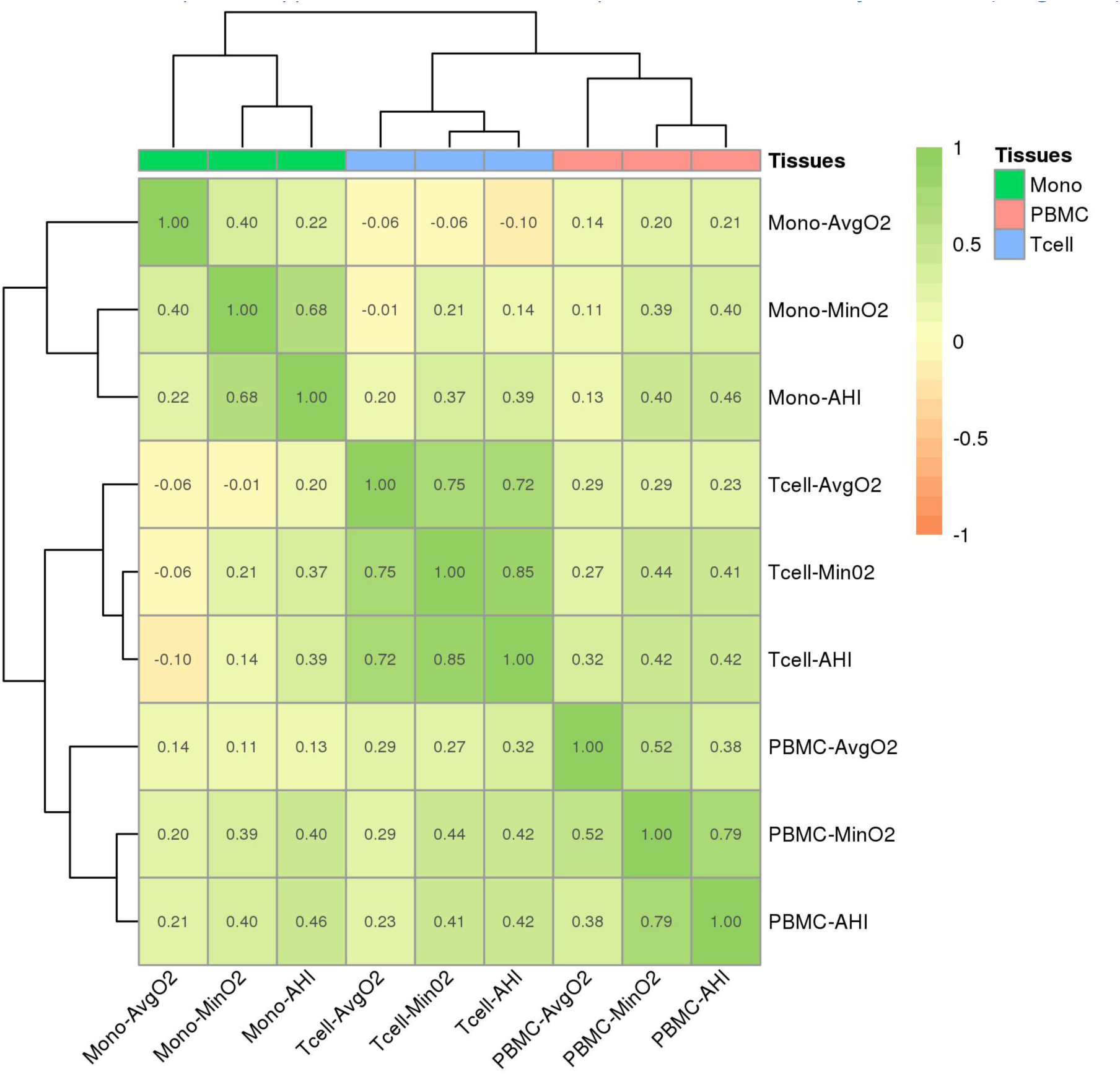
Spearman correlations between estimated log-fold changes in gene expression across SDB phenotypes and tissues in analysis without BMI adjustment (all genes). Heatmap illustrating the Spearman correlations of log-fold change of transcript expression by tissue type (monocytes, T-cells, PBMCs) and SDB phenotype (AvgO2, MinO2, AHI). Color legend portrays Spearman R^2^ (no/weak correlation = light yellow; complete/strong correlation = green).

**Figure S8:**
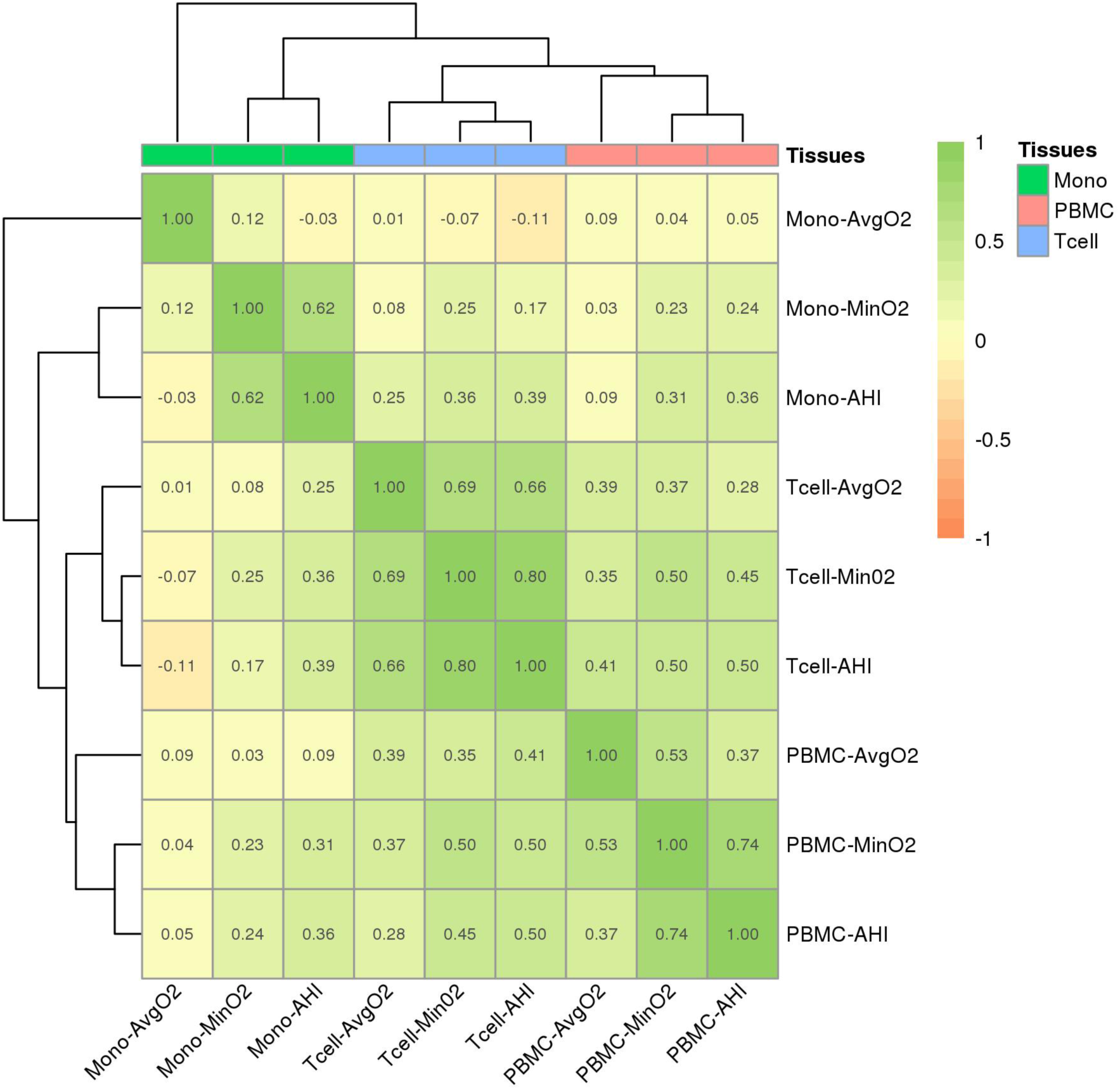
Spearman correlations between estimated log-fold changes in gene expression across SDB phenotypes and tissues in analysis BMI adjustment (all genes). Heatmap illustrating the Spearman correlations of log-fold change of transcript expression by tissue type (monocytes, T-cells, PBMCs) and SDB phenotype (AvgO2, MinO2, AHI). Color legend portrays Spearman R^2^ (no/weak correlation = light yellow; complete/strong correlation = green).

**Figure S9.**
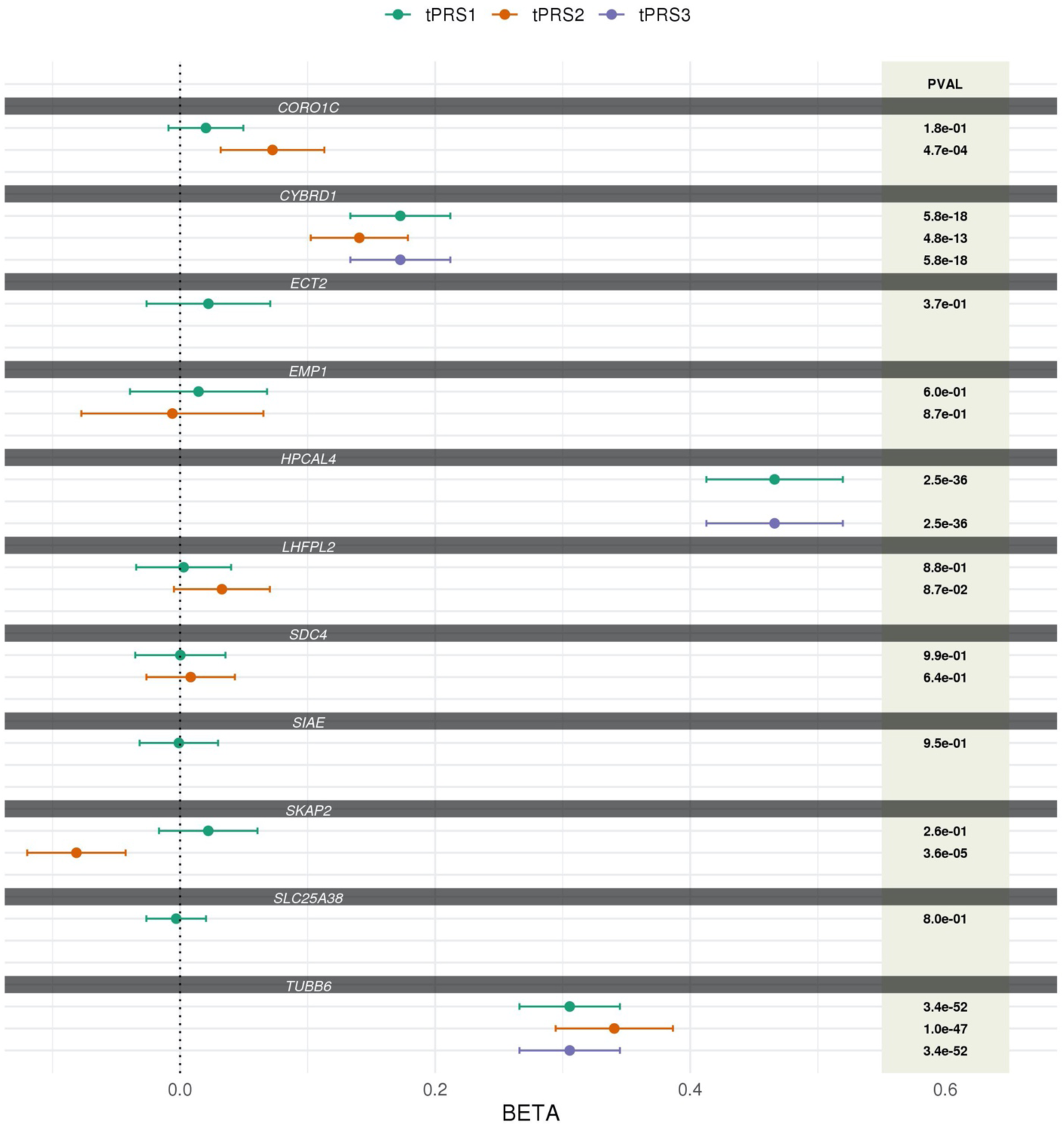
Comparison of the associations between monocyte-based tPRSs and whole-blood gene expression in WHI. For each transcript associated with an SDB phenotype, the figure provides the estimated association effect, 95% confidence interval, and p-value, of tPRSs constructed in different approaches with whole-blood transcript expression in WHI. tPRS1 and tPRS3 were constructed using the clump and threshold approach implemented in PRSice2 using summary statistics from GWAS of transcript expression in monocytes in MESA, and with clumping guided by LD in MESA (the same individuals used for GWAS). tPRS1 allows for genome-wide SNPs, and tPRS3 focused on cis-eQTLs. tPRS2 is the prediXcan model (also using cis-eQLTs only). For tPRS1 and 3 we considered three p-value threshold (5x10^-8^, 10^-7^, and 10^-6^), and the tPRS with smallest p-value is displayed. PRS associations were estimated in models adjusted for sex, age, study site, race/ethnic background, batch effects, and 11 ancestral principal components.

## Bibliography

1. Rundo JV. Obstructive sleep apnea basics. Cleve Clin J Med. 2019 Sep;86(9 Suppl 1):2–9.

2. Mehra R, Stone KL, Blackwell T, Ancoli Israel S, Dam T-TL, Stefanick ML, et al. Prevalence and correlates of sleep-disordered breathing in older men: osteoporotic fractures in men sleep study. J Am Geriatr Soc. 2007 Sep;55(9):1356–64.

3. Redline S, Kump K, Tishler PV, Browner I, Ferrette V. Gender differences in sleep disordered breathing in a community-based sample. Am J Respir Crit Care Med. 1994 Mar;149(3 Pt 1):722–6.

4. Osorio RS, Gumb T, Pirraglia E, Varga AW, Lu S-E, Lim J, et al. Sleep-disordered breathing advances cognitive decline in the elderly. Neurology. 2015 May 12;84(19):1964– 71.

5. Botros N, Concato J, Mohsenin V, Selim B, Doctor K, Yaggi HK. Obstructive sleep apnea as a risk factor for type 2 diabetes. Am J Med. 2009 Dec;122(12):1122–7.

6. Peppard PE, Young T, Palta M, Skatrud J. Prospective study of the association between sleep-disordered breathing and hypertension. N Engl J Med. 2000 May 11;342(19):1378– 84.

7. Li X, Sotres-Alvarez D, Gallo LC, Ramos AR, Aviles-Santa L, Perreira KM, et al. Associations of Sleep-disordered Breathing and Insomnia with Incident Hypertension and Diabetes. The Hispanic Community Health Study/Study of Latinos. Am J Respir Crit Care Med. 2021 Feb 1;203(3):356–65.

8. Chami HA, Fontes JD, Vasan RS, Keaney JF, O’Connor GT, Larson MG, et al. Vascular inflammation and sleep disordered breathing in a community-based cohort. Sleep. 2013 May 1;36(5):763–768C.

9. Unnikrishnan D, Jun J, Polotsky V. Inflammation in sleep apnea: an update. Rev Endocr Metab Disord. 2015 Mar;16(1):25–34.

10. de Paula LKG, Alvim RO, Pedrosa RP, Horimoto ARVR, Krieger JE, Oliveira CM, et al. Heritability of OSA in a rural population. Chest. 2016 Jan 6;149(1):92–7.

11. Carmelli D, Colrain IM, Swan GE, Bliwise DL. Genetic and environmental influences in sleep-disordered breathing in older male twins. Sleep. 2004 Aug 1;27(5):917–22.

12. Ryan S, Cummins EP, Farre R, Gileles-Hillel A, Jun JC, Oster H, et al. Understanding the pathophysiological mechanisms of cardiometabolic complications in obstructive sleep apnoea: towards personalised treatment approaches. Eur Respir J. 2020 Aug 6;56(2).

13. Baguet J-P, Hammer L, Lévy P, Pierre H, Launois S, Mallion J-M, et al. The severity of oxygen desaturation is predictive of carotid wall thickening and plaque occurrence. Chest. 2005 Nov;128(5):3407–12.

14. Nácher M, Serrano-Mollar A, Farré R, Panés J, Seguí J, Montserrat JM. Recurrent obstructive apneas trigger early systemic inflammation in a rat model of sleep apnea. Respir Physiol Neurobiol. 2007 Jan 15;155(1):93–6.

15. Dematteis M, Godin-Ribuot D, Arnaud C, Ribuot C, Stanke-Labesque F, Pépin J-L, et al. Cardiovascular consequences of sleep-disordered breathing: contribution of animal models to understanding the human disease. ILAR J. 2009;50(3):262–81.

16. Dyugovskaya L, Lavie P, Lavie L. Phenotypic and functional characterization of blood gammadelta T cells in sleep apnea. Am J Respir Crit Care Med. 2003 Jul 15;168(2):242–9.

17. Dyugovskaya L, Lavie P, Lavie L. Increased adhesion molecules expression and production of reactive oxygen species in leukocytes of sleep apnea patients. Am J Respir Crit Care Med. 2002 Apr 1;165(7):934–9.

18. Minoguchi K, Tazaki T, Yokoe T, Minoguchi H, Watanabe Y, Yamamoto M, et al. Elevated production of tumor necrosis factor-alpha by monocytes in patients with obstructive sleep apnea syndrome. Chest. 2004 Nov;126(5):1473–9.

19. Bergeron C, Kimoff J, Hamid Q. Obstructive sleep apnea syndrome and inflammation. J Allergy Clin Immunol. 2005 Dec;116(6):1393–6.

20. Gharib SA, Seiger AN, Hayes AL, Mehra R, Patel SR. Treatment of obstructive sleep apnea alters cancer-associated transcriptional signatures in circulating leukocytes. Sleep. 2014 Apr 1;37(4):709–14, 714A.

21. Perry JC, Guindalini C, Bittencourt L, Garbuio S, Mazzotti DR, Tufik S. Whole blood hypoxia-related gene expression reveals novel pathways to obstructive sleep apnea in humans. Respir Physiol Neurobiol. 2013 Dec 1;189(3):649–54.

22. Turnbull CD, Lee LYW, Starkey T, Sen D, Stradling J, Petousi N. Transcriptomics Identify a Unique Intermittent Hypoxia-mediated Profile in Obstructive Sleep Apnea. Am J Respir Crit Care Med. 2020 Jan 15;201(2):247–50.

23. Sofer T, Li R, Joehanes R, Lin H, Gower AC, Wang H, et al. Transcriptional survey of peripheral blood links lower oxygen saturation during sleep with reduced expressions of CD1D and RAB20 that is reversed by CPAP therapy. medRxiv. 2019 Aug 12;

24. Polotsky VY, Bevans-Fonti S, Grigoryev DN, Punjabi NM. Intermittent hypoxia alters gene expression in peripheral blood mononuclear cells of healthy volunteers. PLoS One. 2015 Dec 14;10(12):e0144725.

25. Alterki A, Joseph S, Thanaraj TA, Al-Khairi I, Cherian P, Channanath A, et al. Targeted Metabolomics Analysis on Obstructive Sleep Apnea Patients after Multilevel Sleep Surgery. Metabolites. 2020 Sep 1;10(9).

26. Xu H, Zheng X, Jia W, Yin S. Chromatography/Mass Spectrometry-Based Biomarkers in the Field of Obstructive Sleep Apnea. Medicine. 2015 Oct;94(40):e1541.

27. Bild DE, Bluemke DA, Burke GL, Detrano R, Diez Roux AV, Folsom AR, et al. Multi-Ethnic Study of Atherosclerosis: objectives and design. Am J Epidemiol. 2002 Nov 1;156(9):871–81.

28. Chen X, Wang R, Zee P, Lutsey PL, Javaheri S, Alcántara C, et al. Racial/Ethnic Differences in Sleep Disturbances: The Multi-Ethnic Study of Atherosclerosis (MESA). Sleep. 2015 Jun 1;38(6):877–88.

29. Hays J, Hunt JR, Hubbell FA, Anderson GL, Limacher M, Allen C, et al. The Women’s Health Initiative recruitment methods and results. Ann Epidemiol. 2003 Oct;13(9 Suppl):S18-77.

30. Sorlie PD, Avilés-Santa LM, Wassertheil-Smoller S, Kaplan RC, Daviglus ML, Giachello AL, et al. Design and implementation of the Hispanic Community Health Study/Study of Latinos. Ann Epidemiol. 2010 Aug;20(8):629–41.

31. Lavange LM, Kalsbeek WD, Sorlie PD, Avilés-Santa LM, Kaplan RC, Barnhart J, et al. Sample design and cohort selection in the Hispanic Community Health Study/Study of Latinos. Ann Epidemiol. 2010 Aug;20(8):642–9.

32. Conomos MP, Laurie CA, Stilp AM, Gogarten SM, McHugh CP, Nelson SC, et al. Genetic diversity and association studies in US hispanic/latino populations: applications in the hispanic community health study/study of latinos. Am J Hum Genet. 2016 Jan 7;98(1):165–84.

33. Kowalski MH, Qian H, Hou Z, Rosen JD, Tapia AL, Shan Y, et al. Use of >100,000 NHLBI Trans-Omics for Precision Medicine (TOPMed) Consortium whole genome sequences improves imputation quality and detection of rare variant associations in admixed African and Hispanic/Latino populations. PLoS Genet. 2019 Dec 23;15(12):e1008500.

34. Feofanova EV, Chen H, Dai Y, Jia P, Grove ML, Morrison AC, et al. A Genome-wide Association Study Discovers 46 Loci of the Human Metabolome in the Hispanic Community Health Study/Study of Latinos. Am J Hum Genet. 2020 Nov 5;107(5):849–63.

35. Li B, Dewey CN. RSEM: accurate transcript quantification from RNA-Seq data with or without a reference genome. BMC Bioinformatics. 2011 Aug 4;12:323.

36. Evans AM, DeHaven CD, Barrett T, Mitchell M, Milgram E. Integrated, nontargeted ultrahigh performance liquid chromatography/electrospray ionization tandem mass spectrometry platform for the identification and relative quantification of the smallmolecule complement of biological systems. Anal Chem. 2009 Aug 15;81(16):6656–67.

37. Ohta T, Masutomi N, Tsutsui N, Sakairi T, Mitchell M, Milburn MV, et al. Untargeted metabolomic profiling as an evaluative tool of fenofibrate-induced toxicology in Fischer 344 male rats. Toxicol Pathol. 2009 Jun;37(4):521–35.

38. Sofer T, Kurniansyah N, Aguet F, Ardlie K, Durda P, Nickerson DA, et al. Benchmarking association analyses of continuous exposures with RNA-seq in observational studies. Brief Bioinformatics. 2021 May 20;

39. Benjamini Y, Hochberg Y. Controlling the false discovery rate: A practical and powerful approach to multiple testing. Journal of the Royal Statistical Society: Series B (Methodological). 1995 Jan;57(1):289–300.

40. Sofer T, Zheng X, Gogarten SM, Laurie CA, Grinde K, Shaffer JR, et al. A fully adjusted two-stage procedure for rank-normalization in genetic association studies. Genet Epidemiol. 2019 Jan 17;43(3):263–75.

41. Choi SW, O’Reilly PF. PRSice-2: Polygenic Risk Score software for biobank-scale data. Gigascience. 2019 Jul 1;8(7).

42. Gogarten SM, Sofer T, Chen H, Yu C, Brody JA, Thornton TA, et al. Genetic association testing using the GENESIS R/Bioconductor package. Bioinformatics. 2019 Dec 15;35(24):5346–8.

43. Gamazon ER, Wheeler HE, Shah KP, Mozaffari SV, Aquino-Michaels K, Carroll RJ, et al. A gene-based association method for mapping traits using reference transcriptome data. Nat Genet. 2015 Sep;47(9):1091–8.

44. Mogil LS, Andaleon A, Badalamenti A, Dickinson SP, Guo X, Rotter JI, et al. Genetic architecture of gene expression traits across diverse populations. PLoS Genet. 2018 Aug 10;14(8):e1007586.

45. Lumley T. Package “survey.” R package version. 2015;

46. Lumley T. Complex surveys: a guide to analysis using R. 2011;

47. Bridge KS, Sharp TV. Regulators of the hypoxic response: a growing family. Future Oncol. 2012 May;8(5):491–3.

48. Hou Z, Peng H, White DE, Negorev DG, Maul GG, Feng Y, et al. LIM protein Ajuba functions as a nuclear receptor corepressor and negatively regulates retinoic acid signaling. Proc Natl Acad Sci USA. 2010 Feb 16;107(7):2938–43.

49. Ponia SS, Robertson SJ, McNally KL, Subramanian G, Sturdevant GL, Lewis M, et al. Mitophagy antagonism by ZIKV reveals Ajuba as a regulator of PINK1 signaling, PKR-dependent inflammation, and viral invasion of tissues. Cell Rep. 2021 Oct 26;37(4):109888.

50. Reigstad LJ, Varhaug JE, Lillehaug JR. Structural and functional specificities of PDGF-C and PDGF-D, the novel members of the platelet-derived growth factors family. FEBS J. 2005 Nov;272(22):5723–41.

51. Clara CA, Marie SKN, de Almeida JRW, Wakamatsu A, Oba-Shinjo SM, Uno M, et al. Angiogenesis and expression of PDGF-C, VEGF, CD105 and HIF-1α in human glioblastoma. Neuropathology. 2014 Aug;34(4):343–52.

52. Arnardottir ES, Nikonova EV, Shockley KR, Podtelezhnikov AA, Anafi RC, Tanis KQ, et al. Blood-gene expression reveals reduced circadian rhythmicity in individuals resistant to sleep deprivation. Sleep. 2014 Oct 1;37(10):1589–600.

53. Chen A, Sceneay J, Gödde N, Kinwel T, Ham S, Thompson EW, et al. Intermittent hypoxia induces a metastatic phenotype in breast cancer. Oncogene. 2018 Aug;37(31):4214–25.

54. Monard D. SERPINE2/Protease Nexin-1 in vivo multiple functions: Does the puzzle make sense? Semin Cell Dev Biol. 2017 Feb;62:160–9.

55. Demeo DL, Mariani TJ, Lange C, Srisuma S, Litonjua AA, Celedon JC, et al. The SERPINE2 gene is associated with chronic obstructive pulmonary disease. Am J Hum Genet. 2006 Feb;78(2):253–64.

56. Chappell S, Daly L, Morgan K, Baranes TG. The SERPINE2 gene and chronic obstructive pulmonary disease. American journal of. 2006;

57. Baker JB, Low DA, Simmer RL, Cunningham DD. Protease-nexin: a cellular component that links thrombin and plasminogen activator and mediates their binding to cells. Cell. 1980 Aug;21(1):37–45.

58. Görlach A, Diebold I, Schini-Kerth VB, Berchner-Pfannschmidt U, Roth U, Brandes RP, et al. Thrombin Activates the Hypoxia-Inducible Factor-1 Signaling Pathway in Vascular Smooth Muscle Cells. Circ Res. 2001 Jul 6;89(1):47–54.

59. Solleti SK, Srisuma S, Bhattacharya S, Rangel-Moreno J, Bijli KM, Randall TD, et al. Serpine2 deficiency results in lung lymphocyte accumulation and bronchus-associated lymphoid tissue formation. FASEB J. 2016 Jul;30(7):2615–26.

60. Zolotoff C, Bertoletti L, Gozal D, Mismetti V, Flandrin P, Roche F, et al. Obstructive Sleep Apnea, Hypercoagulability, and the Blood-Brain Barrier. J Clin Med. 2021 Jul 14;10(14).

61. Liak C, Fitzpatrick M. Coagulability in obstructive sleep apnea. Can Respir J. 2011 Dec;18(6):338–48.

62. Garcia-Guzman M, Soto F, Gomez-Hernandez JM, Lund PE, Stühmer W. Characterization of recombinant human P2X4 receptor reveals pharmacological differences to the rat homologue. Mol Pharmacol. 1997 Jan;51(1):109–18.

63. Ozaki T, Muramatsu R, Sasai M, Yamamoto M, Kubota Y, Fujinaka T, et al. The P2X4 receptor is required for neuroprotection via ischemic preconditioning. Sci Rep. 2016 May 13;6:25893.

64. Moore SC, Matthews CE, Sampson JN, Stolzenberg-Solomon RZ, Zheng W, Cai Q, et al. Human metabolic correlates of body mass index. Metabolomics. 2014 Apr 1;10(2):259–69.

65. Bardsley EN, Pen DK, McBryde FD, Ford AP, Paton JFR. The inevitability of ATP as a transmitter in the carotid body. Auton Neurosci. 2021 Sep;234:102815.

66. Zolkipli-Cunningham Z, Naviaux JC, Nakayama T, Hirsch CM, Monk JM, Li K, et al. Metabolic and behavioral features of acute hyperpurinergia and the maternal immune activation mouse model of autism spectrum disorder. PLoS One. 2021 Mar 18;16(3):e0248771.

67. Yang A, Sonin D, Jones L, Barry WH, Liang BT. A beneficial role of cardiac P2X4 receptors in heart failure: rescue of the calsequestrin overexpression model of cardiomyopathy. Am J Physiol Heart Circ Physiol. 2004 Sep;287(3):H1096–103.

68. Hu B, Mei QB, Yao XJ, Smith E, Barry WH, Liang BT. A novel contractile phenotype with cardiac transgenic expression of the human P2X4 receptor. FASEB J. 2001 Dec;15(14):2739–41.

69. Razavi AC, Bazzano LA, He J, Fernandez C, Whelton SP, Krousel-Wood M, et al. Novel findings from a metabolomics study of left ventricular diastolic function: the bogalusa heart study. J Am Heart Assoc. 2020 Feb 4;9(3):e015118.

70. Melis M, Pillolla G, Bisogno T, Minassi A, Petrosino S, Perra S, et al. Protective activation of the endocannabinoid system during ischemia in dopamine neurons. Neurobiol Dis. 2006 Oct;24(1):15–27.

71. Matthews AT, Lee JH, Borazjani A, Mangum LC, Hou X, Ross MK. Oxyradical stress increases the biosynthesis of 2-arachidonoylglycerol: involvement of NADPH oxidase. Am J Physiol Cell Physiol. 2016 Dec 1;311(6):C960–74.

72. von Hinke S, Davey Smith G, Lawlor DA, Propper C, Windmeijer F. Genetic markers as instrumental variables. J Health Econ. 2016 Jan;45:131–48.

73. VanderWeele TJ, Tchetgen Tchetgen EJ, Cornelis M, Kraft P. Methodological challenges in mendelian randomization. Epidemiology. 2014 May;25(3):427–35.

## References

1. Conomos, M.P. et al. (2016) Genetic diversity and association studies in US hispanic/latino populations: applications in the hispanic community health study/study of latinos. Am. J. Hum. Genet. 98, 165–184

2. Kowalski, M.H. et al. (2019) Use of >100,000 NHLBI Trans-Omics for Precision Medicine (TOPMed) Consortium whole genome sequences improves imputation quality and detection of rare variant associations in admixed African and Hispanic/Latino populations. PLoS Genet. 15, e1008500

3. Hays, J. et al. (2003) The Women’s Health Initiative recruitment methods and results. Ann Epidemiol 13, S18–77

